# Cochlear implant-related speech processing may diminish the advantage of exposure to infant-directed speech

**DOI:** 10.1101/2020.06.29.20140319

**Authors:** Meisam K. Arjmandi, Derek Houston, Yuanyuan Wang, Laura C. Dilley

## Abstract

Caregivers modify their speech when talking to infants, a specific type of speech known as infant-directed speech (IDS). This speaking style facilitates language learning compared to adult-directed speech (ADS) in infants with normal hearing (NH). While infants with NH and those with cochlear implants (CIs) prefer listening to IDS over ADS, it is yet unknown how CI speech processing may affect the acoustic distinctiveness between ADS and IDS, as well as the degree of intelligibility of these. This study analyzed speech of seven female adult talkers to model the effects of simulated CI processing on (1) acoustic distinctiveness between ADS and IDS, (2) estimates of intelligibility of caregivers’ speech in ADS and IDS, and (3) individual differences in caregivers’ ADS-to-IDS modification and estimated speech intelligibility. Results suggest that CI processing is substantially detrimental to the acoustic distinctiveness between ADS and IDS, as well as to the intelligibility benefit derived from ADS-to-IDS modifications. Moreover, the observed variability across individual talkers in acoustic implementation of ADS-to-IDS modification and speech intelligibility was significantly reduced due to CI processing. The findings are discussed in the context of the link between IDS and language learning in infants with CIs.

## I. INTRODUCTION

### A. Infant-directed speech and language learning

Caregivers around the world modify their speaking style from adult-directed speech (ADS) to infant-directed speech (IDS) when talking to infants (Fernald & Kuhl, 1987; Snow, 1977). Greater exposure to IDS and some aspects of its acoustic and linguistic properties facilitates language learning in children with normal hearing (NH) (Cristia & Seidl, 2014; Liu, Kuhl, & Tsao, 2003; Song, Demuth, & Morgan, 2010; Trainor, Austin, & Desjardins, 2000; Weisleder & Fernald, 2013). The supportive role of IDS in infants’ language learning occurs through several pathways, such as directing and holding infants’ attention to speech (Fernald, 1985, 1989; Kitamura, Thanavishuth, Burnham, & Luksaneeyanawin, 2001; Kuhl et al., 1997; Schachner & Hannon, 2011; Wang, Bergeson, & Houston, 2017, 2018; Werker, Pegg, & McLeod, 1994), while making speech more clear by providing more salient cues for speech discrimination (Karzon, 1985), word segmentation (Singh & Nestor, 2009), speech segmentation (Thiessen, Hill, & Saffran, 2005), and word learning (Ma, Golinkoff, Houston, & Hirsh-Pasek, 2011). Taken together, these studies demonstrate the supportive role of IDS in infants’ early language learning. However, it is not yet clear how cochlear implants may affect acoustic properties of IDS, and its supportive role in language learning. The present study aims to investigate the effects of simulated CI processing on acoustic information pertaining to distinguishing and recognizing words in IDS versus ADS.

### B. Infants’ attention to IDS

In listening to speech, infants with NH and those with CIs demonstrate increased attention to IDS compared to ADS (The ManyBabies Consortium, 2020; Cooper, Abraham, Berman, & Staska, 1997; Fernald, 1985; Wang et al., 2017; Wang, Bergeson, et al., 2018; Werker & McLeod, 1989; Werker et al., 1994). This effect of IDS on attention begins very early as observed in newborns and infants as young as 4 months old (Cooper & Aslin, 1990; Fernald, 1985; Werker & McLeod, 1989), demonstrating the capability of infants in using subtle acoustic information to distinguish IDS from ADS while having acquired only minimal linguistic knowledge. Multiple acoustic-phonetic cues have been identified as potentially supporting infants’ robust preference for IDS over ADS, such as higher pitch (Cooper & Aslin, 1990; Fernald, 1989), greater pitch fluctuation (Fernald & Simon, 1984), slower speaking rate (Leong, Kalashnikova, Burnham, & Goswami, 2014; Narayan & McDermott, 2016; Song et al., 2010), and an expanded vowel space (Burnham et al., 2015) in IDS compared to ADS. These distinctive acoustic-phonetic cues are manifested in spectro-temporal representations of caregivers’ speech, which are actively incorporated through infants’ highly sensitive auditory systems to recognize IDS as distinct from ADS (Kuhl, 2004; Maye, Werker, & Gerken, 2002; McMurray & Aslin, 2005; Telkemeyer et al., 2009). Although IDS over ADS preference was observed in infants with CIs (Wang et al., 2017), it is still not clear how they are different from their peers with NH in terms of access to spectral information involved in distinguishing IDS from ADS. Infants with CIs may face difficulties in resolving distinctive acoustic information toward performing this task effectively, leading potentially to a disruption in the developmental time course of CI infants’ attention to IDS and delayed language development, as compared to that of NH infants. These considerations suggest a benefit of examining how CI-related speech processing may affect acoustic distinctiveness and intelligibility of IDS compared with ADS, and how this acoustic distinctiveness and intelligibility may differ as a function of individual talkers’ speech patterns.

### C. CI processing and acoustic distinction between IDS and ADS

Infants with CIs have access to a spectro-temporally degraded representation of speech in their language environments, due to limitations of electric stimulation to faithfully transmit this information to the auditory nerve. The limited number of active channels and the broad current spread, that causes the interaction between channels, lead to a degraded representation of spectral and temporal information in speech (Baskent & Gaudrain, 2016; Svirsky, 2017). For instance, Friesen et al. (2001) showed that speech recognition in listeners with NH improved by increasing the number of spectral channels in vocoded speech, highlighting the effect of spectral resolution in recognition of speech in listeners with CIs (Baskent & Gaudrain, 2016; Friesen et al., 2001; Fu, Chinchilla, Nogaki, & Galvin, 2005; Fu & Nogaki, 2005; H. Luo & Poeppel, 2007; Shannon, Zeng, Kamath, Wygonski, & Ekelid, 1995; Svirsky, 2017). Temporal resolution is also reduced in CIs and it is mainly limited to temporal-envelope cues, which disrupts faithful transmission of temporal fine-structure in speech to auditory nerves (Rubinstein, 2004; Svirsky, 2017). This reduced spectro-temporal resolution may negatively affect infants’ access to segmental and suprasegmental (i.e., prosodic) features of IDS.

Transmission of multiple speech cues are negatively affected by the limited resolution of CIs, including pitch (Mehta & Oxenham, 2017; Qin & Oxenham, 2005; Svirsky, 2017), timbre (Kong, Mullangi, Marozeau, & Epstein, 2011), and melody (Mehta & Oxenham, 2017; Zeng, Tang, & Lu, 2014) in talkers’ speech. These cues have been recognized as major attributes that distinguish IDS from ADS (Fernald, 1989; Fernald & Simon, 1984; Wang et al., 2017). For example, caregivers’ pitch is identified as an important perceptual cue for children’s robust attention in listening to IDS (Fernald, 1985; Piazza, Iordan, & Lew-Williams, 2017) and was also recognized as an important cue in distinguishing IDS from ADS (Fernald & Kuhl, 1987). Higher mean pitch and wider pitch range are recognized as prosodic modifications that signal speech directed to infants as compared to adults (Garnica, 1977; Narayan & McDermott, 2016). Talkers’ pitch further contributes to lexical segmentation of speech in listeners with CIs (Spitzer, Liss, Spahr, Dorman, & Lansford, 2009), a task critical for word learning. Recognition of emotion from speech is another important aspect in recognizing and processing IDS (Trainor et al., 2000), which relies heavily on perceiving pitch contour and pitch fluctuation in caregivers’ speech. These cues are poorly perceived by listeners with CIs. For example, listeners with CIs performed worse in emotion recognition as the number of spectral channels in vocoded speech was decreased (Luo, Fu, & Galvin, 2007). Another instance of degraded representation of speech cues related to IDS is poor perception of melodic pitch that is shown to be negatively affected by reducing the number of spectral channels (Kong, Cruz, Jones, & Zeng, 2004). These findings suggest that children with CIs have probably partial access to spectro-temporal cues in speech that greatly contribute to the distinctiveness of IDS and ADS (e.g., pitch and intonation). This limited access to speech cues that signal IDS may interfere with the connection between recognizing and listening to IDS and developing better language skills. Therefore, it is important to understand how CI may impact the acoustic distinction between IDS and ADS - a major knowledge gap that the present study aimed to address based on acoustic and computational analysis of simulated CI speech.

### D. Intelligibility of IDS through CI

Another possible way IDS may foster language learning is by making speech more intelligible. Prior studies showed evidence that IDS may provide acoustic cues that assist infants for parsing linguistic units in speech. Karzon (1985) showed that the exaggerated suprasegmental features of IDS may provide supportive perceptual cues for syllabification of multisyllabic sequences. In another study, infants had better long-term word recognition when the words were presented in IDS compared to ADS (Singh & Nestor, 2009). It was further shown that IDS facilitates word segmentation (Thiessen et al., 2005). Although evidence on intelligibility benefits of IDS over ADS have somehow been contradictory at the level of segmental speech cues (e.g., Kuhl et al., 1997; McMurray, Kovack-Lesh, Goodwin, & McEchron, 2013), there might be an intelligibility benefit associated with other properties of IDS (e.g., prosodic cues), similar to the effects observed in close analogues of a “clear speech” register (Bradlow, Kraus, & Hayes, 2003; S. H. Ferguson & Kewley-Port, 2007). Caregivers’ modification of speaking style from ADS to IDS impacts aspects of speech such as pitch contour and speech rate, which have been shown to be strong predictors of speech clarity and intelligibility (Bradlow et al., 2003; Cutler, Dahan, & van Donselaar, 1997; Ferguson & Poore, 2010; Ferguson & Quené, 2014; Spitzer, Liss, & Mattys, 2007; Watson & Schlauch, 2008). Different choices of speaking style often impact speech rate, where a slower rate is thought to contribute to enhanced intelligibility of clear speech compared to conversational speech both in typical listeners (Ferguson et al., 2010) and recipients of CIs (Li et al., 2011; Zanto, Hennigan, Östberg, Clapp, & Gazzaley, 2013).

Limited spectral resolution of the cochlear implant device affects the quality by which listeners with CIs receive speech cues (Croghan, Duran, & Smith, 2017; Jain & Vipin Ghosh, 2018; Peng, Hess, Saffran, Edwards, & Litovsky, 2019; Qin & Oxenham, 2005), which may negatively impact CI users’ ability to recognize words and phonemes, especially children (Grieco-calub, Saffran, & Litovsky, 2010; Peng et al., 2019). Therefore, any likely beneficial role of IDS in improvement of speech intelligibility is prone to change when caregivers’ speech is perceived through a CI device. Prior studies have shown that performance of listeners with CIs is considerably poorer than listeners with NH in understanding sentences spoken with relatively higher rate (Zanto et al., 2013), which is an acoustic dimension signaling distinction of IDS from ADS (i.e., slower speaking rate in IDS compared to ADS). Variation of fundamental frequency (F_0_) is another acoustic dimension of the IDS-vs-ADS distinction that contributes to speech recognition (Spitzer, Liss, Spahr, Dorman, & Lansford, 2009; Spitzer et al., 2007). Findings on the effect of these acoustic properties on intelligibility of caregivers’ speech is still controversial, and little is known about the effects of CI speech processing on intelligibility of caregivers’ speech in ADS and IDS conditions. The present study uses computational modeling approaches to address whether IDS is beneficial for intelligibility improvement and how this effect may be compromised depending on listeners’ hearing status (NH vs. CI).

### E. Individual differences in acoustic implementation of IDS and speech intelligibility

Listeners’ familiarity with the range of variability in voice of individual talkers is important for robust identification and recognition of individuals’ voices (Lavan, Burton, Scott, & McGettigan, 2019; Souza, Gehani, Wright, & McCloy, 2013). Caregiver-specific acoustic modification is important for effective caregiver-infant interaction and significantly contributes to infants’ recognition of caregivers’ voices both before and after birth (Beauchemin et al., 2011; Kisilevsky et al., 2003), as well as for neural development of infants’ auditory cortices (Webb, Heller, Benson, & Lahav, 2015). Singh (2008) showed that exposure to IDS with relatively higher variability in vocal affects facilitates word recognition in infants by forming a relatively wider and more generalizable lexical category, highlighting the supportive role of exposure to a wide range of ADS-to-IDS acoustic variation in effective processing of caregivers’ speech. The degree of acoustic variability that caregivers introduce to their infants may vary across individuals. The acoustic characteristics of IDS reveal finer details about the unique voice timbre of individual caregivers, which potentially contributes to infant’ identification of individual caregivers (Piazza et al., 2017). It is, however, unclear how CI speech processing impacts the range of acoustic variability within and across caregivers due to shifts in caregivers’ speaking style (i.e., ADS to IDS), a change which likely eliminates acoustic details and potentially negatively impacts the supportive role of IDS in recognizing caregivers’ voices. CI speech processing is also detrimental to the acoustic distance of voices across caregivers, which potentially makes distinguishing talkers from each other more challenging for infants with CIs, compared to those with NH. The present study aimed to investigate these questions by focusing on simulating the limited resolution of CIs using noise-excited envelope vocoder and modeling the intelligibility benefit of ADS-to-IDS modification.

### F. Benefits of investigations of acoustic properties of CI-related speech processing

Studies aimed at understanding speech perception in individuals with CIs can be characterized as involving one of three general approaches. The first approach examines performance of listeners with NH in response to vocoded speech to simulate hearing in listeners with CIs (Dorman, Loizou, & Rainey, 1997a; Jahn, DiNino, & Arenberg, 2019; Mehta & Oxenham, 2017; Qin & Oxenham, 2003, 2005). The second method involves directly study of how listeners with CIs perform in various speech recognition tasks (Brown & Bacon, 2010; Kong, Stickney, & Zeng, 2005; Peng, Tomblin, & Turner, 2008; Peng, Hess, Saffran, Edwards, & Litovsky, 2019). In a third method – the one which we focus on in the present paper – acoustic properties of simulated CI speech and unprocessed analogs are analyzed comparatively. This comparative analysis can be facilitated through the use of various quantitative metrics that emulate speech recognition in CI users as proxies to analyze properties of CI speech and its perception in listeners with CIs (Jain & Vipin Ghosh, 2018; Qin & Oxenham, 2003; Santos, Cosentino, Hazrati, Loizou, & Falk, 2013). Analysis of simulated CI speech and quantitative measures of speech quality and intelligibility tailored to listeners with CIs provide repeatable, automated, inexpensive, and fast tools for gaining preliminary evidence about speech perception in individuals with CIs. As such, a comparative analysis method offers benefits for undertaking efficient investigations for further study, and as such provides benefits over various major challenges that accompany the first two categories of studies, such as participant recruitment, time, and cost.

### G. Current study

In the present study, we analyzed unprocessed and simulated CI speech of seven female talkers who spoke fifteen utterances both in ADS and IDS. These pairs of ADS and IDS speech were analyzed to examine how the limited frequency resolution of CIs may affect acoustic distinctiveness between ADS and IDS, as well as its effects on the estimated intelligibility of caregivers’ speech under modifications of speaking style (i.e., ADS to IDS). ADS and IDS stimuli of the seven female talkers were processed using noise-excited envelope vocoder with different number of channels to simulate the restricted spectral resolution and the amount of distortion in CIs (Dorman, Loizou, Fitzke, & Tu, 1998; Dorman, Loizou, & Rainey, 1997b; Friesen et al., 2001; Fu, Shannon, & Wang, 1998; Loizou, Dorman, & Tu, 1999; Mehta, Lu, & Oxenham, 2020; Mehta & Oxenham, 2017; Qin & Oxenham, 2003, 2005; Rosen, Faulkner, & Wilkinson, 1999; Shannon et al., 1995).

To examine the effects of CIs on acoustic distinctiveness between IDS and ADS, we first calculated mel-frequency cepstral coefficients (MFCCs), which have been shown in multiple studies to be effective for characterizing the distinctive qualities of IDS vs. ADS (Inoue, Nakagawa, Kondou, Koga, & Shinohara, 2011; Piazza et al., 2017; Sulpizio et al., 2018). Modeling the frequency response of human auditory system via MFCCs captures acoustic features that are involved in distinguishing IDS and ADS, such as shifts in vocal timbre (Piazza et al., 2017) and acoustic distinctiveness between IDS and ADS (Inoue et al., 2011), including in other languages (e.g., Italian and German) (Sulpizio et al., 2018). MFCCs are also able to effectively reflect caregivers’ emotional state and vocal affect (e.g., happiness vs. sadness, Sato & Obuchi, 2007; Slaney & McRoberts, 1998), attributes that help explain infants’ preference for IDS over ADS (Fernald, 2018; Horowitz, 1983; Mastropieri & Turkewitz, 1999; Moore, Spence, & Katz, 1997; Papoušek, Bornstein, Nuzzo, Papoušek, & Symmes, 1990; Singh, Morgan, & Best, 2002; Walker-Andrews & Grolnick, 1983; Walker-Andrews & Lennon, 1991). Furthermore, MFCCs allowed us to analyze spectral properties of vocoded speech, which was not feasible to do based on calculation of common acoustic properties such as fundamental frequency (F_0_), as these cues are partially present in CI-simulated speech (Fuller et al., 2014; Gaudrain & Baskent, 2018). Second, to quantify the acoustic distinctiveness between IDS and ADS, we calculated a *Mahalanobis distance (MD)* measure over MFCCs features. MD is a multivariate distance metric that has been widely used to measure the distances between vectors in a variety of multidimensional feature spaces (Arjmandi, Dilley, & Wagner, 2018; Masnan et al., 2015; Xiang, Nie, & Zhang, 2008). Third, to model how CIs may influence intelligibility of caregivers’ speech, we calculated the speech-to-reverberation-modulation energy ratio (SRMR) to model the signal-based intelligibility of speech signals delivered to listeners with NH, as well as its CI-tailored version (SRMR-CI) to model intelligibility of speech signals delivered to listeners with CIs (Santos et al., 2013). These measures were examined both within and across speakers to model the impacts of speaking styles (IDS vs. ADS) and listener group (NH vs. CI) on signals’ distinctiveness and estimated intelligibility. All these analyses were implemented in Matlab 2019a (The Mathworks Web-Site [http://www.mathworks.com]).

The following four specific questions were addressed in this study. First, we asked how simulated CI speech processing affects the acoustic distinctiveness of IDS compared with ADS, particularly as a function of degree of spectral degradation (i.e., number of spectral channels) in CI-simulated speech. We hypothesized that (a) CI-related speech processing significantly degrades the acoustic distinctiveness of IDS compared with ADS, and further that (b) increasing the number of channels would not compensate for degradation imposed by CI-related processing as compared to the unprocessed condition. Second, we asked whether there is signal-based evidence that IDS may be more intelligible than ADS, as gauged by the SRMR metric (to simulate a NH listening condition) or the SRMR-CI metric (to simulate a CI listening condition). We hypothesized that IDS is more intelligible than ADS, but that estimated intelligibility would vary as a function of simulated listening status (NH vs. CI). Third, we asked to what extent the acoustic distinctiveness between IDS and ADS varies across individual caregivers, as well as how such individual differences might be impacted by CI speech processing, focusing on change in the spectral resolution. We predicted individual differences across caregivers in acoustic implementation of the differences between ADS and IDS; we further predicted that CI-related speech processing would decrease the extent of acoustic distinctiveness as a function of speech style, leading to loss of intra- and inter-subject acoustic variability in terms of IDS vs. ADS distinctiveness. Fourth, and finally, we asked how estimated intelligibility *differences* for IDS compared with ADS vary across individual caregivers, and how such intelligibility differ as a function of hearing status (as gauged by SRMR vs. SRMR-CI differences to simulate effects of NH vs. CI listening conditions, respectively). We hypothesized that intelligibility differences for IDS vs. ADS would vary across caregivers, and that these intelligibility differences would be more similar as estimated for CI listeners, compared with NH listeners.

## II. METHODS

### A. Speech stimuli

Seven female adult native talkers of American English ranging in age from 21 to 24 years old spoke fifteen utterances in IDS and ADS (See Appendix in Supplementary Material for the list of stimuli). The utterances were elicited to be also used in separate infant word-learning experiments in which infants were exposed to a novel target word (i.e., “*modi*”) in the context of behavioral measures of infant word recognition to assess whether infants learned the novel word from IDS better than ADS. To elicit stimuli, the talkers were instructed to speak utterances as if talking to an infant (IDS condition) or an adult (ADS condition); this procedure was similar to that used in Wang et al., (2017; 2018).

Speech stimuli were recorded using an AKG D 542 ST-S microphone in a sound booth and digitized at a sampling rate of 44.1 kHz with 16-bit resolution. The distance between talkers’ mouths and the microphone was controlled to assure the quality of the recorded stimuli. Prior to processing as discussed below, the start and end points of recorded utterances were manually identified in Praat software (Boersma & Weenink, 2001) to remove preceding and following silent portions. All participants were fully informed about the purpose and procedure of this study, and they had given informed consent to participate. This study was approved by the Institutional Review Boards of the Ohio State University and Michigan State University.

### B. Creation of simulated CI speech

CI-simulated versions of the unprocessed stimuli were created using noise-excited envelope vocoder processing at six levels of spectral degradation, corresponding to 4-, 8-, 12-, 16-, 22- and 32-channel noise-vocoded stimuli. The choice of number of channels was made to cover both the actual number of channels in FDA-approved devices (12, 16, 22) and to query ranges of variation ranging from minimal cues used in prior studies (4, 8) out to 32 channels to examine CI vocoder scenarios with higher spectral resolution. The natural stimuli consisting of spoken IDS or ADS were processed in AngelSim^™^ Cochlear Implant and Hearing Loss Simulator (Fu, 2019; Emily Shannon Fu Foundation, www.tigerspeech.com) using 4-, 8-, 12-, 16-, 22-, and 32-channel noise-vocoding CI-simulated stimuli; the noise vocoding method followed the procedure in Shannon et al. (1995). The original stimuli were first band-passed filtered using the Greenwood function (emulating the Greenwood frequency-place map) into *N* (*N* = {4, 8, 12, 16, 22, 32}) adjacent frequency channels ranging from 200 Hz to 8000 Hz. This was implemented in *AngelSim*^*™*^ by setting the absolute lower- and higher-frequency threshold for analysis and carrier filters to 200 Hz and 8000 Hz with a filter slope of 24 dB/Oct (Fu, 2019). These frequency ranges are fairly close to the corner frequencies of the Cochlear Nucleus speech processors in CI listeners cochlear implant devices (Crew & Galvin, 2012; Winn & Litovsky, 2015), which emulate the performance of average CI listeners in speech envelope discrimination (Chatterjee & Oberzut, 2011; Chatterjee & Peng, 2008). The same analysis filter and carrier filter of the Greenwood function was used to analyze white noise as a carrier signal (Greenwood, 1990). The *AngelSim* software used this setup to extract a time-varying amplitude envelope of speech stimuli under each frequency band using half-wave rectification and then modulated independent white-noise carriers. There were 210 stimuli per speech style condition (7 talkers x 15 stimuli x 2 speaking styles). Including the noise-vocoded versions of these utterances (at 4, 8, 12, 16, 22, and 32-channels), we analyzed 1470 (210 x 7 levels of speech degradation) utterances in the present study.

### C. Using MFCC features to characterize acoustic properties of IDS and ADS

We calculated 12 MFCCs for each speech stimulus to characterize its acoustic information (Hunt, Lennig, & Mermeletein, 1980; Imai, 1983; Shaneh & Taheri, 2009). Figure 1 illustrates this step for a sample pair of IDS-ADS stimuli in the process for measuring the acoustic distinctiveness between each ADS-IDS stimulus pair. To calculate MFCCs, each stimulus was re-sampled at 16 kHz using a Hamming window of 25 ms applied to each frame, with a frame shift of 10 ms, following the procedure used in Inoue et al. (2011). For each pair of IDS and ADS stimuli (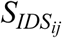 and 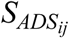 in Figure 1), MFCCs were calculated for all frames of these stimuli (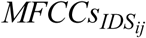 and 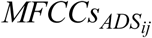). Here, *i* indicates the index of the speech stimulus, *i* = {1,2,3,…,15}, and *j* is the index for the talker, *j =*{1,2,3,…,7}. Unlike previous studies in which a single, time-averaged MFCC vector was calculated to represent acoustic information in IDS and ADS (e.g., Piazza et al., 2017), our approach took advantage of all MFCCs derived from all frames of a speech stimuli to calculate the acoustic distinctiveness between ADS-IDS pairs of stimuli within a multidimensional feature space, thereby preserving the details about spectro-temporal information in speech at the level of frame. Since each analyzed pair of IDS and ADS stimuli contained identical word strings, our analysis was expected to mainly model the acoustic effects of caregivers’ speaking styles (IDS vs. ADS).

**FIG. 1.**
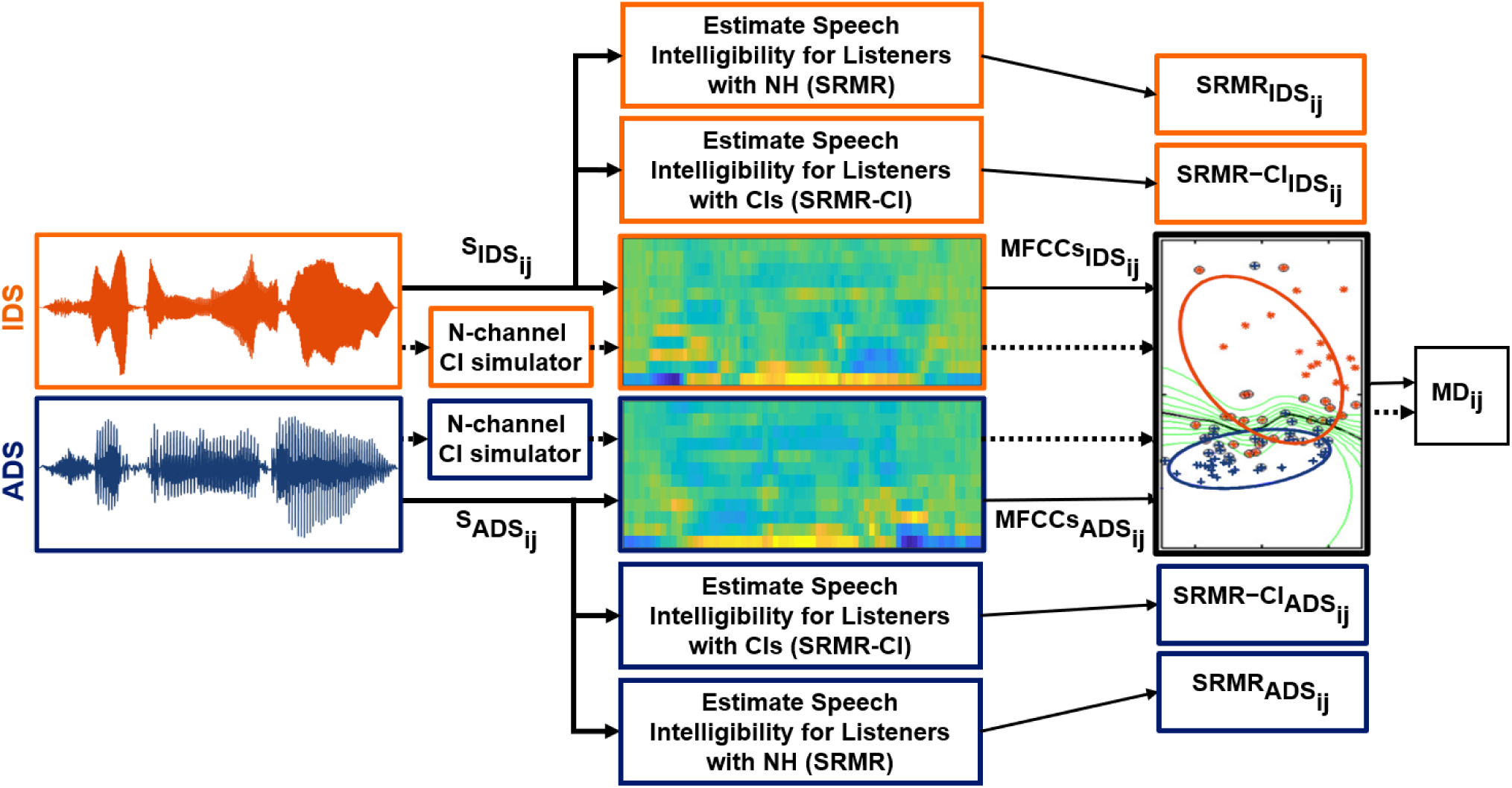
Schematic diagram of the approach used in the present study for measuring (1) MD between pairs of MFCCs derived from pairs of IDS-ADS stimuli, and (2) intelligibility of IDS and ADS stimuli as estimated by SRMR value for listeners with NH and SRMR-CI for those with CIs. Note that the dashed line denotes the process for creating and analyzing the noise-vocoded versions of the same pairs of stimuli, while N stands for the number of spectral channels in the noise-excited envelope vocoder. Blocks and lines with blue (dark gray) color indicate paths for processing ADS, while those with orange (light gray) color indicate paths for processing IDS. Note that SRMR and SRMR-CI were calculated only for unprocessed stimuli in order to estimate intelligibility for NH and CI listeners, respectively. In this figure, the waveforms and their corresponding MFCCs are from the utterance “See the modi?” spoken by one of the seven talkers both in IDS and ADS speaking styles. 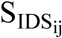 and 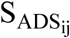 are the i^th^ pair of IDS and ADS stimuli (i = {1,2,3,…,15}) for talker j (j ={1,2,3,…,7}). 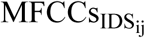 and 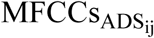 are MFCC features derived from 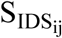 and 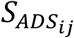 speech stimuli, respectively. The middle two panels show MFCCs obtained from the frames of these IDS and ADS stimuli. MD_ij_ is the MD calculated to measure the acoustic distance between the two matrices for 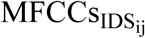 and 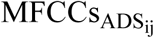. 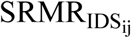 and 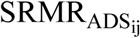 are the estimated intelligibility for 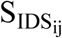 and 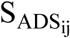 speech stimuli, respectively, as heard by listeners with NH. 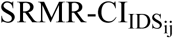 and 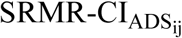 are the estimated speech intelligibility for the same 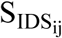 and 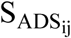 speech stimuli, respectively, as heard by a listener with CIs.

### D. Acoustic distinctiveness quantification using Mahalanobis Distance (MD) measure

After representing acoustic features of pairs of IDS and ADS stimuli by calculating their MFCCs, the acoustic distinctiveness between each pair was measured by calculating MD on the corresponding MFCC matrices (i.e 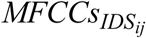 and 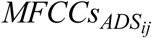) using a 12-dimensional feature space (i.e., 12 MFCCs) (Maesschalck & Massart, 2000; Masnan et al., 2015; Xiang et al., 2008). MD is a multivariate statistical approach that evaluates distances between two multidimensional feature vectors or matrices that belong to two classes (here IDS vs. ADS) (Arjmandi et al., 2018; Heijden, Ferdinand, Ridder, & Tax, 2005; Maesschalck & Massart, 2000; Masnan et al., 2015). The MD calculation returns the *distance* between means of two classes (here IDS and ADS) relative to the average per-class covariance matrix (Maesschalck & Massart, 2000). Here, acoustic properties of each class were represented by two feature matrices (here, MFCCs). The larger the distance between pairs of 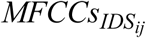 and 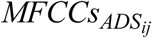 feature vectors, the lower the overlap between two classes of IDS and ADS; this corresponds in turn to greater acoustic distance (or distinctiveness) between IDS and ADS stimuli.

Each row of the calculated MFCCs matrix for utterance *i* from talker *j* (e.g., 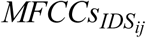 or 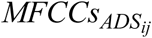) corresponds to a frame of that utterance, and 12 MFCCs were presented on the columns of this matrix. Therefore, the dimension of each MFCC matrix was *N*_*F*_ x 12, where *N*_*F*_ refers to the number of frames in that speech stimulus. The acoustic distinctiveness between IDS and ADS stimuli for each female talker *j* was computed by averaging the fifteen MD values obtained from that talker’s fifteen IDS-ADS pairs 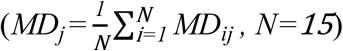. This process was separately performed on pairs of IDS and ADS stimuli for seven levels of spectral degradation – *unprocessed, 32-, 22-, 16-, 12-, 8-*, and/or *4-channel* noise-vocoded CI-simulated stimuli – to give average MDs used to examine (1) how the acoustic distinctiveness between IDS and ADS stimuli changes as a function of CI processing (and number of spectral channels), (2) how individual caregivers vary in acoustic distinctiveness between IDS and ADS stimuli, and (3) how the limited spectral resolution in CIs may affect this pattern of individual differences across talker.

### E. Estimation of speech intelligibility using quantitative metrics of speech-to-reverberation-modulation energy ratio (SRMR)

As shown in Figure 1, we estimated the degree of intelligibility of each speech stimulus as it might be experienced by listeners with NH or with CIs. To approximate stimulus intelligibility as associated with NH, we calculated the quantitative metric of *speech-to-reverberation-modulation energy ratio* (SRMR) for IDS and ADS (unprocessed) stimuli, respectively (Santos et al., 2013). This metric has previously been validated as an approximation of intelligibility of speech to listeners with NH in behavioral tasks (Falk et al., 2015; Santos et al., 2013). Further, to approximate intelligibility as might be experienced by listeners with CIs, we calculated an adapted version of the SRMR metric, which was also developed by Falk et al. (2013) to tailor the SRMR metric for CI users; this adapted metric is known as SRMR-CI (Santos et al., 2013).

To calculate SRMR, an (unprocessed) speech stimulus was first filtered by a 23-channel gammatone filterbank in order to emulate cochlear function. Next, a Hilbert transform was applied to output signals from each of the 23 filters in the filterbank to obtain their temporal envelopes. Next, modulation spectral energy for each critical band was calculated by windowing stimulus temporal envelope and computing the discrete Fourier transform. To emulate frequency selectivity in the modulation domain, the modulation frequency bins were grouped into eight overlapping modulation bands with logarithmically-spaced center frequencies between 4 and 128 Hz. Finally, the SRMR value was obtained by calculating the ratio of the average modulation energy in the first four modulation bands (∼ 3–20 Hz) to the average modulation energy in the last four modulation bands (∼ 20-160 Hz), as explained in Santos et al., (2013).

To calculate SRMR-CI, a 22-channel filterbank was used instead of a 23-channel filterbank in order to emulate the structure of filterbanks in Nucleus CI devices. In addition, the 4-128 Hz range of modulation for filterbank center frequencies was replaced by a 4–64 Hz range to better emulate CI users’ performance (Santos et al., 2013). Although these measures have not primarily been developed to estimate the effect of talkers’ speaking style on speech intelligibility, the signal processing algorithms used in these metrics estimate the distribution of energy in various frequency bands, as well as other spectral properties that are expected to emulate fairly well performance of NH and CI users in speech recognition. Intelligibility of signals as estimated for different hearing statuses (NH or CI) is inherently modeled through the difference in algorithmic implementation of these metrics (SRMR vs. SRMR-CI, respectively) (Santos et al., 2013).

As shown in Figure 1, SRMR and SRMR-CI values were separately calculated (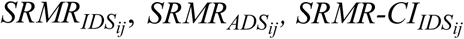 and 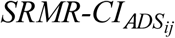) for each pair of stimuli in IDS and ADS conditions (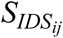 and 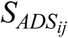). Intelligibility of speech stimuli produced by talker *j* in the two conditions (ADS and IDS) was summarized by averaging the relevant SRMR-related values over the 15 stimuli spoken by this talker at each of these two conditions. Therefore, for talker *j*, four values were calculated: (i) 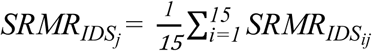, (ii) 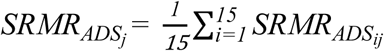, (iii) 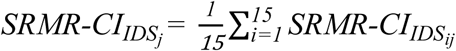, (iv) 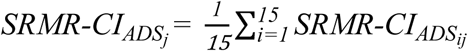.These four values for each talker *j* provided an approximation of the intelligibility of IDS and ADS stimuli spoken by this talker, as heard by listeners with NH (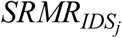 and 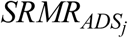) and by those with CIs (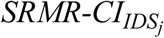 and 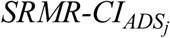). These values were then used to address (1) how the intelligibility of speech is affected by the two separate factors of caregivers’ speaking styles (ADS vs. IDS), as well as by (simulated) group hearing status (NH vs. CI), (2) the extent to which the degree of impact of ADS-to-IDS speaking style modifications on intelligibility varies across individual caregivers, and (3) how this variation is affected by (simulated) hearing status (NH vs. CI).

### F. Statistical analysis

We constructed two Generalized Linear Mixed Models (GLMMs) in Matlab (using the *fitglme* function) (Matuschek, Kliegl, Vasishth, Baayen, & Bates, 2017; Quené & van den Bergh, 2008) in order to (1) identify whether changes in acoustic distinctiveness (i.e., MD) between IDS and ADS due to CI noise vocoding were statistically significant, and (2) to examine the effect of speaking style (IDS vs. ADS) and simulated hearing status (NH vs. CI), and any potential interaction, on intelligibility of speech (as measured by SRMR and SRMR-CI to simulate NH and CI hearing statuses, respectively). For the first GLMM, the speech degradation level (i.e., 4, 8, 12, 16, 22, and 32-channel, and unprocessed) was entered into the model as a fixed predictor, and MDs were defined as the response variable. The individual female talkers and speech stimuli were defined as random effects (intercepts) in the model to account for quality differences that might exist due to talker- and stimulus-specific variation. A post hoc test using Tukey’s multiple comparison approach (*multcompare*, MATLAB, Mathworks) was conducted to examine statistically significant mean differences for all possible pairwise comparisons across seven levels of spectral degradation (4, 8, 12, 16, 22, 32-channels, and unprocessed). In the second GLMM, factors of speaking style (IDS or ADS) and simulated hearing status (NH or CI) were entered into the model as independent fixed variables; the estimated speech intelligibility (cf. SRMR or SRMR-CI values) corresponded to the response variable in the model. Talkers and speech stimuli were entered as random-effect intercepts in the model to account for quality and/or information differences that might exist due to talker- and stimulus-specific variation.

Measures of central tendency and variability (i.e., mean, standard deviation, and range) were calculated as descriptive statistics to evaluate the degree of variability across caregivers in acoustic implementation of IDS and ADS. We further calculated coefficients of variance (CVs) for MD differences between ADS and IDS conditions for both unprocessed and simulated CI speech within a 22-channel noise-vocoder to examine whether variability across individual caregivers in ADS-IDS acoustic distinctiveness decreased as a function of speech degradation (unprocessed vs. 22-channel simulated CI speech). The same analysis was performed to examine whether changes in speech intelligibility – due to an ADS-to-IDS style shift – were variable across talkers and/or how variability changed as (simulated) listener hearing status changed (from NH to CI). We further calculated Pearson correlation coefficients to examine whether changes in acoustic distinctiveness and speech intelligibility due to changes in speaking style (ADS to IDS) across degradation levels (unprocessed vs. 22-channel vocoded) and hearing status (NH vs. CI) involved a linear transformation.

## III. RESULTS

### B. Effects of CI-related spectral degradation on ADS-vs-IDS acoustic distinctiveness

Fig. 2 shows the mean and distribution of MDs obtained as a function of signal degradation that ranged from a no-degradation (i.e., unprocessed) condition to CI-simulated speech for which the number of spectral channels in the noise-vocoder was gradually reduced from 32 to 4. Overall, MDs between IDS and ADS monotonically decreased with decreasing numbers of spectral channels in the noise-vocoder, thereby highlighting the detrimental effects of limited spectral resolution in CI devices for signal fidelity. As illustrated in Fig. 2, the acoustic distinctiveness between IDS and ADS is reduced by CI speech processing systems and further reduced through decreasing the number of spectral channels. Even the highest-fidelity CI-simulated condition (32-channel noise-vocoded speech) showed a substantial drop in acoustic distinctiveness between IDS and ADS, relative to the unprocessed condition. The average MD across the six noise-vocoded conditions showed a decline of approximately 67% relative to MD at the unprocessed condition, indicating that a large portion of acoustic information involved in conveying the distinction between IDS and ADS was lost due to CI-related speech processing.

**FIG. 2.**
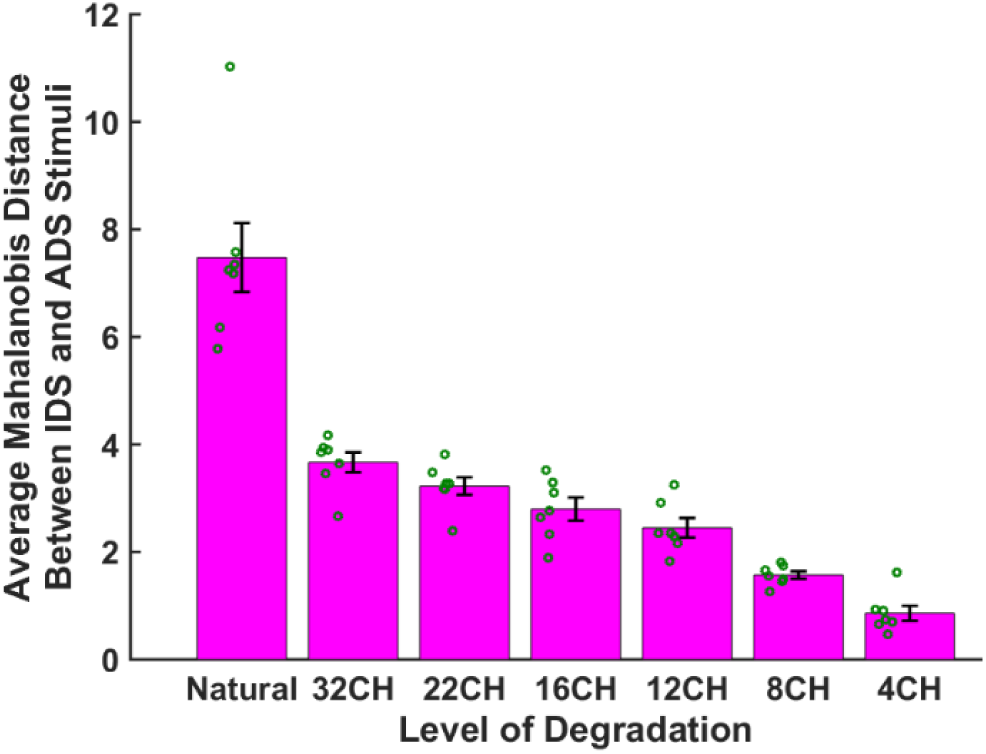
The mean (bar) and ±1 standard error (vertical error bar in black) of Mahalanobis distance (MD) across groups of IDS and ADS stimuli at seven levels of spectro-temporal degradation, ranging from no-degradation (natural/unprocessed) to 4-channel noise-vocoded stimuli. Green (dark gray) circles show the mean MDs for each talker derived by averaging MDs over 15 pairs of IDS-ADS stimuli for that talker.

Table I represents the results of the GLMM analysis to test statistical significance of the effect of CI-related speech processing on MD. As this table suggests, the detrimental effect of CI-related processing, as captured in the decrease in the number of spectral channels on the distinctiveness between IDS and ADS was statistically significant (*β* = −0.886, *t* = −16.54, *p* < 0.0001). Post hoc testing using a Tukey’s multiple comparison approach revealed that the simulated effect of limited spectral resolution in CI processing on the decrease in MDs was statistically significant for comparisons of all pairs of speech degradation conditions except three (32 vs. 22 channels, 22 vs. 16 channels, and 16 vs. 12 channels).

**TABLE I.** Statistical GLMM for modeling the effect of level of speech degradation on acoustic distinctiveness between IDS and ADS as quantified by MD.

| | $\beta$ Estimate | St. Error | t | Pr(>\|t\|) |
| --- | --- | --- | --- | --- |
| (intercept) | 6.694 | 0.58 | 11.45 | 0.000<sup>a</sup> |
| Level of degradation | -0.886 | 0.053 | -16.54 | 0.000<sup>a</sup> |

<sup>a</sup>The p-value was statistically significant.

### B. Effects of CI-related speech processing on intelligibility of ADS and IDS

Fig. 3 shows average SRMR or SRMR-CI values estimating intelligibility of ADS and IDS stimuli for NH and CI listeners. The bar graphs show the group data, corresponding to the means averaged across talkers. The figure suggests that speech spoken in IDS is more intelligible than matched utterances in ADS, regardless of whether intelligibility is estimated for listeners with NH or those with CIs. On average, the stimuli spoken in IDS were ∼15% more intelligible than those spoken in ADS for listeners with normal hearing, as measured by the SRMR metric. This average improvement in intelligibility of IDS stimuli over ADS stimuli was ∼5.7% for listeners with CIs, as estimated by SRMR-CI, suggesting that the supportive effects of IDS for speech intelligibility improvement is decreased through CIs.

**FIG. 3.**
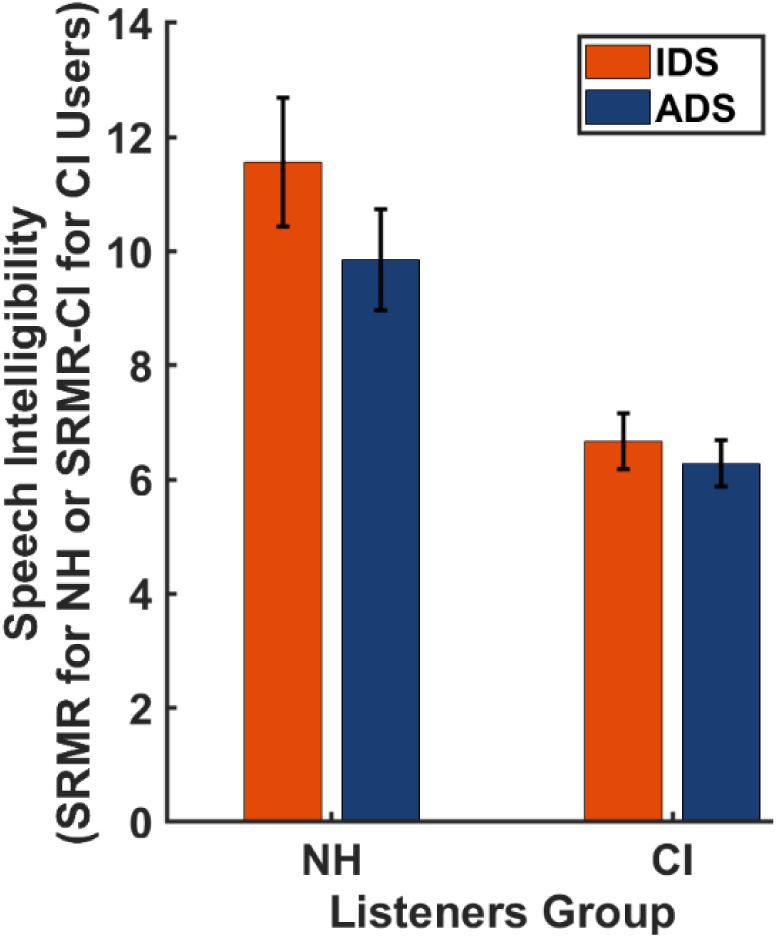
Estimated intelligibility of speech stimuli spoken in IDS (orange; light gray) and in ADS (blue; dark gray) styles (simulated) groups of listeners with different hearing statuses: NH (estimated by SRMR) and those with CIs (estimated by SRMR-CI). The bar graphs represent average values of SRMR or SRMR-CI over the seven talkers.

The results from the GLMM analysis for examining the effect of talkers’ speaking style (IDS vs. ADS), listeners’ group (NH vs. CI), and their interaction on intelligibility of speech (Table II) revealed a significant effect of speaking style (*β* = 3.05, *SE* = 0.74, *t* = 4.14, *p* < 0.0001). This analysis provides new evidence that IDS may likely improve intelligibility of caregivers’ speech to both listeners with NH and those with CIs. These results further showed a significant, negative effect of CI-related processing on intelligibility of caregivers’ speech (*β* = −2.23, *SE* = 0.74, *t* = - 3.02, *p* < 0.0001). In addition, a significant interactive effect between speaking style and listeners group was revealed by the GLMM model (*β* = −1.337, *SE* = 0.466, *t* = −2.865, *p* < 0.001), reflecting the fact that hearing speech through CIs resulted in a smaller intelligibility gain from IDS relative to ADS than was observed in the NH condition.

**TABLE II.**
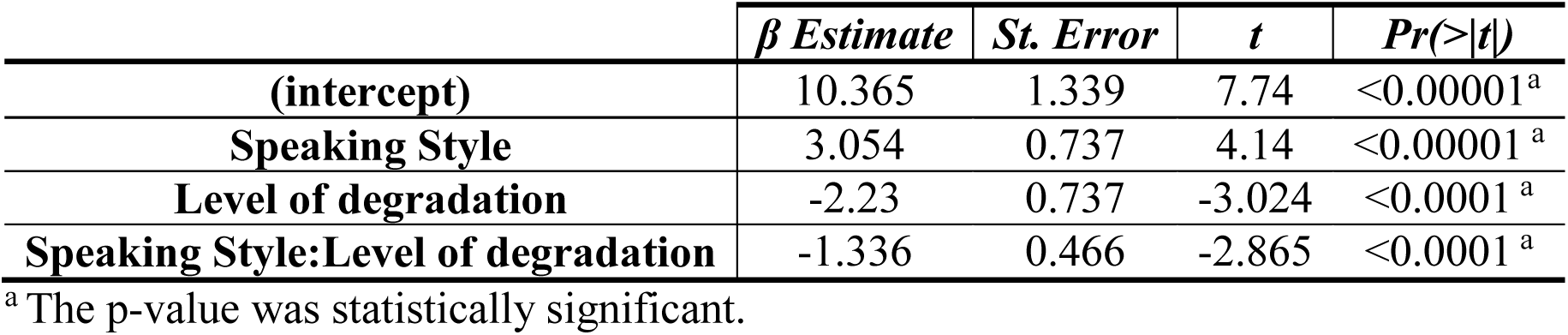
Statistical GLMM for the effects of speaking style, level of speech degradation, and their interaction on intelligibility of speech, as measured by SRMR or SRMR-CI.

### C. Individual differences across caregivers in ADS-vs-IDS acoustic distinctiveness and the effect of simulated CI processing

To examine individual differences across talkers in acoustic distinctiveness of IDS from ADS, we focused on MD data from the individual talkers in two conditions (unprocessed and 22-channel CI-simulated noise-vocoded speech), as shown in Fig. 4A (left vs. right bars, respectively). Two general observations are noticeable in this data. First, caregivers vary considerably in terms of the amount of acoustic variability created by their ADS-to-IDS modification (mean = 7.47, *SD* = 1.70, range = 5.23). For example, the ADS-to-IDS shift in MD for talker 2 exposes a child with NH (cf. natural, unprocessed condition) to a much wider range of acoustic information compared to that for talker 3. This, in turn, suggests that a child of talker 2 would be more advantaged in learning acoustic patterns of her caregivers’ speech compared to a child of talker 3.

**FIG. 4.**
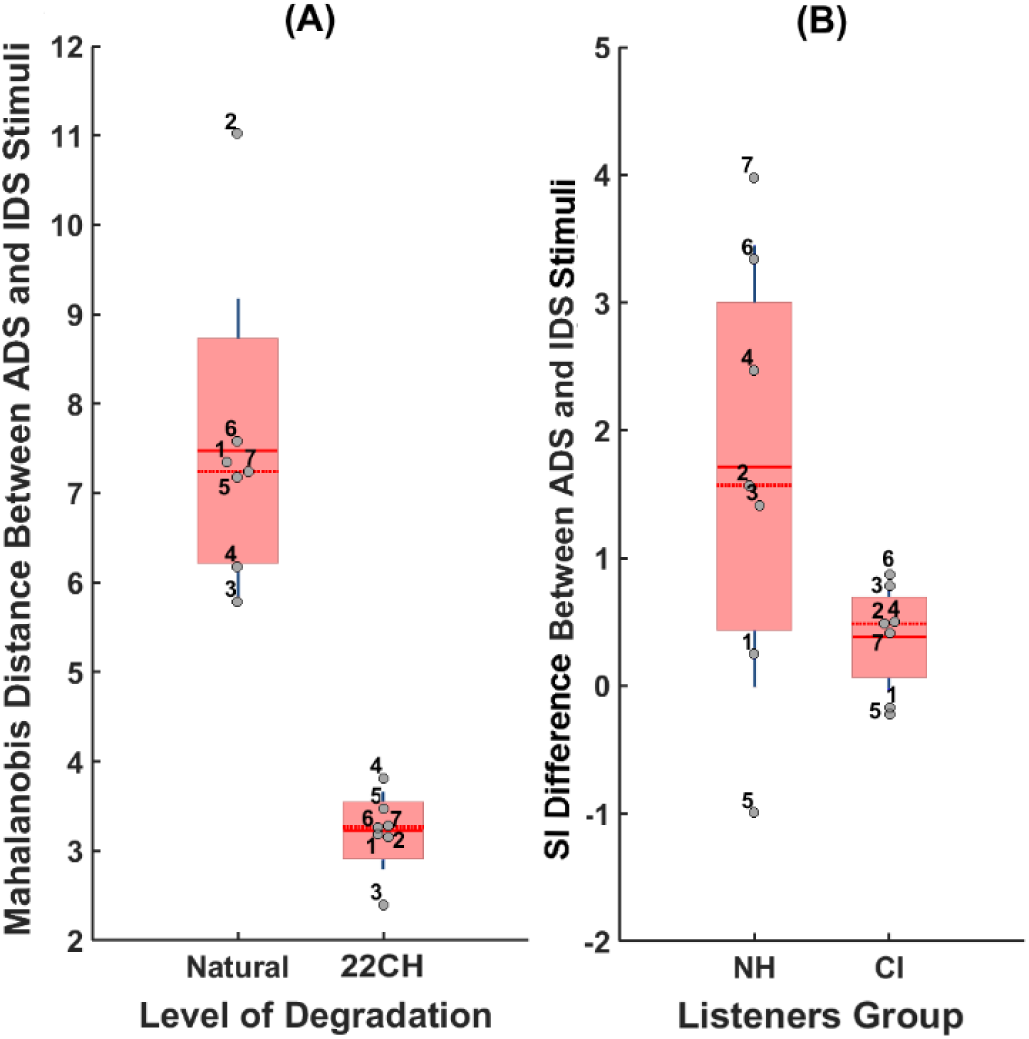
Variability across seven female talkers in (A) acoustic distinctiveness between their IDS and ADS for unprocessed stimuli (Natural) and simulated CI speech within a 22-channel noise vocoder (22CH), and (B) the change in talkers’ speech intelligibility (SI) due to a change in their speaking style (ADS to IDS), as heard for two (simulated) listener groups with either NH (estimated by SRMR) or CIs (estimated by SRMR-CI). The data points (gray circles) are laid over a 1.96 standard error of the mean (95% confidence interval) in red (rectangle area with light gray) and 1 standard deviation shown by blue lines (vertical dark gray lines). The solid and dotted red lines (horizontal solid and dotted dark gray lines) show the mean and median, respectively.

The second observation is that the magnitude of this acoustic variability significantly reduced for CI-simulated signals (mean = 3.22, SD = 0.43, range = 1.4). An analysis of coefficient of variation (CV) revealed that the standard deviation of MD for unprocessed (natural) speech stimuli was 22.7% of its mean, whereas this value was 13.3% for CI-simulated speech stimuli for the 22-channel noise-vocoder. Comparing these two CV values shows that the magnitude of variation in ADS-IDS acoustic distinctiveness (i.e., MD) reduces approximately by half due to CI-related speech processing, highlighting a major loss of acoustic information useful for recognition of caregivers themselves and for distinguishing them from other caregivers.

If CI-simulated processing involves a *linear* transformation of acoustic distinctiveness across speech styles for each talker, then we should observe MD differences to be significantly correlated across natural (i.e., unprocessed) and 22-channel noise-vocoded conditions. The Pearson’s correlation coefficient was *r*(5) = 0.07 (*p* = 0.87), suggesting that CI-simulated speech processing did not impact talkers’ speech proportionately in terms of the acoustic distinctiveness between their IDS and ADS. Instead, CI-related speech processing reduced the IDS-ADS acoustic distance for some talkers (e.g., talker 6) more than others (e.g., talker 4).

### D. Individual differences across caregivers in IDS and ADS intelligibility and the effect of hearing status

To examine how individual talkers vary in the effects of their ADS-to-IDS modifications on intelligibility of speech to listeners with NH and those with CIs, we focused on patterns of SRMR and SRMR-CI metrics from the individual talkers. For (simulated) listeners with NH, SRMRs of pairs of IDS and ADS stimuli were subtracted for each talker and averaged over 15 pairs of ADS-IDS stimuli in order to measure the amount of change in her speech intelligibility due to ADS-to-IDS modification (for talker *j*, 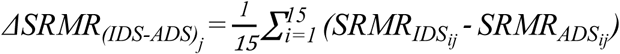). Likewise, for (simulated) listeners with CI, SRMR-CIs of pairs of IDS and ADS stimuli were subtracted for each talker and averaged over 15 pairs of ADS-IDS stimuli in order to quantify the amount of change in her speech intelligibility due to ADS-to-IDS modification (for talker *j*, 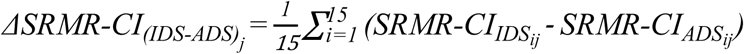). difference measure represents IDS being, on average, more intelligible than ADS for a talker, showing an intelligibility benefit for IDS over ADS for that talker.

Figure 4B (right panel) shows the results for changes in intelligibility of speech of talkers due to modifications to speaking style. The bar plot on the left in Figure 4B (labeled as NH on the x-axis) presents the difference across seven talkers in IDS-ADS intelligibility as estimated for listeners with NH. The data in this scatter plot shows how much speech of a talker (e.g., talker 6) would be heard as more or less intelligible as the talker speaks the utterances or words in IDS compared to ADS.

Two patterns are observable in Figure 4B. First, the change in intelligibility of talkers’ speech due to the speaking style modification (ADS to IDS) was fairly variable across talkers for simulated listeners with NH (mean = 1.71, *SD* = 1.73, range = 4.97), such that this change was associated with relatively more intelligible speech for some talkers compared to others. As this plot suggests, when talkers change their speaking style from ADS to IDS, not only does this not lead to an equal amount of change in intelligibility of speech across talkers, but for one talker the direction of this effect is even slightly negative (talker 5). For the majority of talkers (6 out of 7 talkers), IDS was more intelligible than ADS (as shown by a positive difference value).

Second, relative to the size of the ADS-to-IDS shift for simulated NH listeners, the size of this ADS-to-IDS shift was overall considerably reduced for simulated listeners with CIs (mean = 0.38, SD = 0.43, range = 1.10). The bar plot on the right panel of figure 4B (labeled as CI on the x-axis) presents the dispersion of change in speech intelligibility for each talker due to changing speaking style from ADS to IDS, as estimated for listeners with CIs. Notably, CI-related processing decreases the degree of variability in speech intelligibility, compared with that for NH, as expected. An analysis of CV showed that although the mean of change in speech intelligibility due to ADS-to-IDS modifications was overall smaller for the simulated CI condition compared to the simulated NH condition, while the magnitude of variability was almost the same for two groups of listeners (CV ≅11%). Highlighting non-linearity of effects of CI processing on speech across different talkers, there was no significant correlation across the seven talkers for ADS-to-IDS intelligibility differences in simulated CI vs. NH listening conditions (*r*(5) = 0.74, *p* = 0.58).

## IV. DISCUSSIONS

The present study investigated the simulated effects of CI speech processing on acoustic distinctiveness and intelligibility of speech signals as a function of IDS and ADS speaking styles. Results from the present study supported the hypothesis that the limited spectral resolution in CIs significantly degrades acoustic information involved in distinguishing IDS from ADS. This significant loss of acoustic information related to the distinction between IDS and ADS may negatively impact infants’ recognizing IDS as distinct from ADS, leading possibly to less attention to caregivers’ speech, with potential consequences for children with CIs developing relatively poorer language skills compared to those with NH.

These results are in line with prior findings demonstrating difficulties by listeners with CIs in distinguishing among various speaking styles (Tamati, Janse, & Başkent, 2019). Distinguishing caregivers’ speaking styles is tied to advances in language development (Karzon, 1985; Singh & Nestor, 2009; Thiessen et al., 2005; Wang et al., 2017), but is made challenging when speech is degraded by CIs and/or other undesirable sources, e.g., noise and reverberation (Fetterman & Domico, 2002; Hazrati & Loizou, 2012; Zheng, Koehnke, & Besing, 2011). The significant elimination of IDS-related acoustic information suggested by the present study indicates that infants with CIs likely do not have access to as wide a range of spectro-temporal information in caregivers’ speech to foster recognizing when speech is directed to them. As a result, these children may experience considerable difficulties in recognizing IDS as distinct from ADS, particularly in complex linguistic environments. It should be noted that, in the absence of fine-grained spectral cues, infants with CIs may utilize other well-coded cues through CI devices such as caregivers’ speaking rate to detect whether speech is directed to them.

Although Wang et al., (2017) showed that IDS enhanced attention to speech in infants with CIs compared to ADS, results from the present study suggest that infants’ capability to recognize IDS from ADS and to prefer attending to IDS over ADS is not probably comparable to that of children with NH and is expected to be largely reduced. Furthermore, the fact that infants with CIs showed this preference after 12 months of CI experience in Wang et al.’s (2017) study highlights the possibility that the time course of developing certain capabilities for differentiating IDS and ADS would be longer than peers with NH and would depend on the amount of experience with CIs.

Our results also suggest that infants with CIs must develop a different cue-weighting system for recognition of IDS from ADS compared to infants with NH, where the type and magnitude of the relevant acoustic cues would be different from what infants with NH develop. For example, in the absence of prominent acoustic cues of IDS such as F_0_ (Fernald & Mazzie, 1991; Kuhl & Meltzoff, 1999; Mehler, 1981), which is poorly perceived through CI devices (Mehta et al., 2020; Mehta & Oxenham, 2017; Qin & Oxenham, 2003), infants with CIs may rely more on suprasegmental cues such as speech rate (Peng et al., 2017) and/or periodicity in the temporal envelope (Fu, Chinchilla, & Galvin, 2004; Kong et al., 2004). However, perception of pitch through periodicity in the temporal envelope is mostly limited to F_0_s below around 300 Hz (Carlyon, Deeks, & McKay, 2010; Kong & Carlyon, 2010), which is generally below the range of F_0_ variation in IDS. This lack of access to the entire spectro-temporal cue range in IDS puts infants with CIs at high risk for missing IDS-related communicative events and thus for developing sub-optimal language skills.

Our results corroborated our hypothesis that caregivers likely expose infants to more intelligible speech when speaking in IDS compared to ADS, providing the first evidence on the intelligibility benefit of IDS over ADS. This was shown using a novel application of a recently-developed metric of intelligibility, which models intelligibility of speech for listeners with NH (i.e., SRMR) and those with CIs (i.e., SRMR-CI) (Falk et al., 2015; Santos et al., 2013). This positive effect of IDS on speech intelligibility might be because caregivers provide more clear speech by speaking louder, slower, and/or in a hyperarticulated fashion when using IDS (Hazan et al., 2018; Krause & Braida, 2002; Li et al., 2011; Liu, Del Rio, Bradlow, & Zeng, 2004). These specific speaking patterns likely lead to an intelligibility benefit compared to ADS (Janse, Nooteboom, & Quené, 2007; Liu et al., 2004). Greater intelligibility for IDS over ADS suggests that use of IDS during caregiver-infant spoken communication assists infants in better understanding speech, conceivably supporting infants’ word learning process through a direct link between exposure to IDS and improved speech intelligibility. However, our results also revealed an interactive effect, showing that the size of this positive effect of IDS on caregivers’ speech intelligibility is smaller for infants with CIs compared to their peers with NH. This, in turn, suggests that the limited spectral resolution of CI devices not only disrupts the communication of IDS-ADS-specific acoustic information, but also decreases the intelligibility of caregivers’ IDS, indicating that infants with CIs are not expected to benefit from exposure to IDS as much as their NH peers. These results are consistent with prior findings demonstrating greater difficulties that adult listeners with CIs incur in processing speech and accomplishing lexical access, compared to listeners with NH (McMurray, Farris-Trimble, & Rigler, 2017; Nagels, Bastiaanse, Başkent, & Wagner, 2020). Although prior studies demonstrated a supportive role of IDS on infants’ language outcomes, to our knowledge, this is the first study that provides evidence supporting detrimental effects of the CI speech processing on the supportive role of IDS in speech understanding.

Prior studies on speech recognition in listeners with CIs have highlighted the importance of further examining patterns of individual differences, in addition to group differences (Dilley et al., 2020; Nagels et al., 2020; Peng et al., 2019; Spencer, 2004; Szagun & Schramm, 2016). Our investigation of patterns of individual differences across caregivers in acoustic distinctiveness between IDS and ADS and the simulated effect of limited spectral resolution in CI on these patterns highlighted multiple important observations. The first observation relates to the amount of acoustic variability produced by each talker due to changing speaking style from ADS to IDS. The amount of resultant acoustic variability varied across talkers, indicating that infants would experience different language environments in terms of qualities of their caregivers’ speech. This is expected to result in some caregivers’ exposing infants to relatively larger ranges of acoustic information, as compared to others, which would foster their infants’ speech processing and language acquisition by assisting infants in more robust recognition of their caregivers’ voices (Beauchemin et al., 2011; Kisilevsky et al., 2003), as well as better development of auditory cortical processing for language development (Webb et al., 2015). Similar to the recent findings by Dilley et al. (2020), these results also suggest that IDS is not always readily distinguishable from ADS, due to the fact that caregivers vary in implementation of IDS. As such, these results confirm that conditions for recognition of IDS by infants is not always optimal (Piazza et al., 2017) and might differentially affect language outcomes for infants with CIs (Dilley et al., 2020).

More importantly, the amount of ADS-IDS acoustic difference was considerably reduced for each caregiver as her speech passed through a CI simulator, indicating that infants with CIs probably have access to a much narrower range of acoustic variability in the voices of their caregivers, as compared to their peers with NH. This large decline in the degree of acoustic variability in caregivers’ voices may negatively impact infants’ robust identification of their caregivers’ voices, thereby preventing their gaining the maximal benefit from language input that may be available in their linguistic environments. In fact, encoding and learning these cues is crucial as they both significantly contribute to infants’ familiarity with ranges of acoustic variation in talkers’ speech and their robust performance in understanding speech, despite of multiple sources of variability, such as talker variation (Allen, Miller, & DeSteno, 2009; Eskenazi, 1993), language context (Mattys, 2000; McMurray & Aslin, 2005; Miller, 1994), speech rate (Sommers, Nygaard, & Pisoni, 1992), and background noise and/or reverberation (Hawkins, 2004). Thus, infants with CIs are probably at risk for partial learning of subtle voice cues specific to their caregivers, as reflected in their fine-structure spectro-temporal information.

Acoustic implementation of ADS-to-IDS modification varies across individual caregivers, which may differentially impact infants’ understanding of speech and thus their language outcomes (Dilley et al., 2020; Hoff, 2006; Weisleder & Fernald, 2013). Results from the present study showed that individual talkers varied in the effect of their speaking style modification on speech intelligibility, suggesting that modification of speaking style from ADS to IDS does not always cause an equal degree of improvement in intelligibility of caregivers’ speech. Notably, our results showed that this variability across caregivers in the impact of their speaking style modification on speech intelligibility was reduced due to limited spectral resolution in CIs. In the present study, we only studied variability across caregivers in acoustic information and intelligibility of their IDS compared to ADS, whereas caregivers of infants with CIs may vary in other aspects of spoken communication such as gestural and proprioceptive behaviors, which very likely change the degree of intelligibility by which infants eventually perceive caregivers’ speech (Kirk et al., 2007; Kirk & Pisoni, 2002; Lachs, Pisoni, & Kirk, 2001).

These findings can be further interpreted in the context of spectro-temporal information available to infants’ auditory systems, which is very sensitive to subtle changes in speech acoustics (Jusczyk, Hohne, & Bauman, 1999; Kuhl, 2004). When infants’ accessibility to fine-grained spectro-temporal structures in speech is largely compromised because of limited spectral resolution in CIs, they may not be able to readily recognize and attend to rich IDS. In the absence of this fine-structure spectral information at the output of CI electrodes, infants must rely on course-grained cues in the speech envelope, such as temporal envelope periodicity (Green, Faulkner, & Rosen, 2002; Moore, 2003) or cues from other sensory modalities (e.g., visual and tactile, Green, Nip, Wilson, Mefferd, & Yunusova, 2010; Rohlfing, Fritsch, Wrede, & Jungmann, 2006) in order to recognize IDS from ADS. This increases the cognitive load in processing speech particularly in complex auditory environments and negatively contributes to the observed poor language outcomes in some children (Davidson, Geers, & Nicholas, 2014; Dunn et al., 2014; Geers, Strube, Tobey, & Moog, 2011; Geers, Nicholas, Tobey, & Davidsonb, 2015; Houston, Pisoni, Kirk, Ying, & Miyamoto, 2003; Houston, Stewart, Moberly, Hollich, & Miyamoto, 2012; Miyamoto, Houston, Kirk, Perdew, & Svirsky, 2003; Niparko et al., 2010; Pisoni et al., 2007; Pisoni, Kronenberger, Harris, & Moberly, 2018).

The present study used a within-talker manipulation of speech style to investigate how limited spectral resolution in CIs may affect processing of speech style shifts which are tied to language development, namely IDS vs. ADS signals. While the study involved a sizeable number of individual utterances with control of phonetic properties within talkers, the amount of speech collected from each of the seven talkers was relatively small. Further work will be needed to test how these results extend to a larger sample of talkers with utterances with more varied segmental and lexical composition. Furthermore, studying natural IDS and ADS would be ideal; however, it is difficult to control the content and quality of the speech collected from naturalistic environments. Importantly, studies based on simulated CI speech and computational models of human hearing provide valuable evidence for understanding speech perception in listeners with normal and impaired hearing (Litvak, Delgutte, & Eddington, 2001; Mehta et al., 2020; Rubinstein, Wilson, Finley, & Abbas, 1999; Throckmorton & Collins, 2002). Findings from these studies should be viewed as evidence for general trends, rather than the actual performance of listeners with CIs, subject to further investigation. Although we simulated limitation of CIs in spectral resolution by changing the number of channels in noise-vocoded speech, the effect of other aspects of CI processing on spectral resolution, such as channel interactions (i.e., steepness of spectral slope) (Crew & Galvin, 2012; Mehta et al., 2020), requires further investigation. Additionally, metrics of speech intelligibility of SRMR and SRMR-CI have been validated for adult listeners with CIs and are not direct estimates of speech intelligibility, which is very likely different from what infants with CIs experience in terms of recognition of utterances. Considering these limitations, the results should not be taken as the final determination of how children with CIs perform in recognition between IDS and ADS and how ADS-to-IDS modification impacts the degree of intelligibility of caregivers’ speech to children with CIs.

Despite these limitations, the simulated results from the present study imply that, compared to infants with NH, infants with CIs could be disadvantaged in perceiving IDS and acquiring spoken language due to multiple factors. First, partial transmission of spectro-temporal cues because of limited spectral resolution likely decreases the supportive role of IDS in infants’ language learning by disrupting the link between attending to IDS and speech comprehension. In fact, degraded representation of spectral information that contributes to acoustic distinction between IDS and ADS in CIs may have detrimental effects on infants’ ability to pay attention to caregivers’ speech, something that is a fundamental cognitive skill for spoken language acquisition (Bergeson, 2014; Glenn, Cunningham, & Joyce, 1981; Houston et al., 2003; Rottmann & Zobrist, 2004; Wang, Shafto, & Houston, 2018). It is worth mentioning that infants’ language learning involves incorporating information from multiple interwind communication dimensions (i.e., visual, social, tactile, and emotional), which creates a very rich channel for learning language through infant-directed speech (Gogate, Bahrick, & Watson, 2000; Nomikou & Rohlfing, 2011), even in the absence of major spectro-temporal cues such as talker’s F_0_ in the output of CI electrodes. Our findings further suggested that CIs diminish the benefit of exposure to more intelligible IDS, as compared to ADS, which may negatively impact infants’ abilities to process caregivers’ speech, possibly with further significant downstream consequences for later language skills. Last but not least, our simulated results imply that, compared to infants with NH, infants with CIs have access to a relatively narrower range of acoustic information (corresponding to smaller acoustic variability) in their caregivers’ speech, which probably leads to experiencing greater difficulties in robust identification and recognition of their caregivers’ voices (Beauchemin et al., 2011; Kisilevsky et al., 2003; Lavan et al., 2019), as well as developing poorer word recognition skills (Singh, 2008) and an impaired auditory system for language processing (Webb et al., 2015). Despite the reduced ADS-vs-IDS distinctiveness, intelligibility, and variability of vocoded IDS, it is possible that children with CIs may have developed certain coping/adapting strategies to mitigate the degraded speech input. For example, it is possible that children with CIs may have higher sensitivity (lower threshold) to the acoustic cues than children with NH.

In summary, the current study used computational and signal processing approaches in order to provide new evidence for how CI-related speech processing may impact recognition of IDS from ADS in children with CIs, as well as how these style differences may affect intelligibility benefits derived by style shifts from ADS to IDS. These findings provide solid grounding for developing new perceptual studies to test abilities of infants with CIs to recognize their caregivers’ speaking style (ADS vs. IDS) and to recognize intelligible words in caregivers’ speech. Focusing on computational metrics as undertaken here provides an important complement to costly, complex, labor-intensive perceptual studies. The results provided preliminary evidence for how CI-related speech processing may alter the pathway from exposure to IDS to processing speech and, by extension, the acquisition of language by children with CIs compared to that of their NH peers in two ways: (1) making it probably harder for children with CIs to recognize IDS from ADS, (2) decreasing the ADS-to-IDS intelligibility benefit. The most direct and immediate implication of these findings is the imperative need to improve signal processing in CI devices to assure the faithful transmission of acoustic cues relevant to identification and recognition of IDS from ADS. Until then, the major clinical implication of these findings is that the maximum benefit from exposure to IDS for language learning in infants with CIs requires caregivers’ active use of multimodal (i.e., gesture, tactile, visual, social, and emotional) communicative behaviors in order to compensate for the degraded representation of acoustic information relevant to IDS and to support its robust perception.

## Data Availability

The data in this study will be available upon request.

## ACKNOWLEDGMENTS

Research reported in this publication was supported by the National Institute on Deafness and other Communicative Disorders of the National Institutes of Health under award number R01DC008581 to D. Houston and L. Dilley.

## REFERENCES

Allen, J. S., Miller, J. L., & DeSteno, D. (2009). Individual talker differences in voice-onset-time: Contextual influences. The Journal of the Acoustical Society of America, 125(6), 3974–3982.

Arjmandi, M., Dilley, L. C., & Wagner, S. E. (2018). Investigation of acoustic dimension use in dialect production: machine learning of sonorant sounds for modeling acoustic cues of African American dialect. 11th International Conference on Voice Physiology and Biomechanics, 12–13. East Lansing, USA.

Baskent, D., & Gaudrain, E. (2016). Perception and Psychoacoustics of Speech in Cochlear Implant Users. Scientific Foundations of Audiology. Perspectives from Physics, Biology, Modelling, and Medicine, 185–320. Retrieved from https://books.google.de/books?hl=de&lr=&id=EtAyDAAAQBAJ&oi=fnd&pg=PA285&dq=Scientific+Foundations+of+Audiology&ots=cfEfTicv7h&sig=1cTQmXsc_FR7oNQiwWYgpklOkN0

Beauchemin, M., González-Frankenberger, B., Tremblay, J., Vannasing, P., Martínez-Montes, E., Belin, P., … Lassonde, M. (2011). Mother and stranger: An electrophysiological study of voice processing in newborns. Cerebral Cortex, 21(8), 1705–1711.

Bergeson, T. R. (2014). Hearing versus Listening: Attention to Speech and Its Role in Language Acquisition in Deaf Infants with Cochlear Implants. Lingua, 10–25.

Boersma, P., & Weenink, D. (2001). Praat, a system for doing phonetics by computer. Glot International, 5:9/10, 341–345.

Bradlow, A. R., Kraus, N., & Hayes, E. (2003). Speaking clearly for children with learning disabilities: Sentence perception in noise. Journal of Speech, Language, and Hearing Research, 46(1), 80–97.

Brown, C. A., & Bacon, S. P. (2010). Fundamental frequency and speech intelligibility in background noise. Hearing Research, 266(1–2), 52–59.

Burnham, E. B., Wieland, E. A., Kondaurova, M. V., McAuley, J. D., Bergeson, T. R., & Dilley, L. C. (2015). Phonetic Modification of Vowel Space in Storybook Speech to Infants up to 2 Years of Age. Journal of Speech, Language, and Hearing Research, 58(2), 241–253.

Carlyon, R. P., Deeks, J. M., & McKay, C. M. (2010). The upper limit of temporal pitch for cochlear-implant listeners: Stimulus duration, conditioner pulses, and the number of electrodes stimulated. The Journal of the Acoustical Society of America, 127(3), 1469–1478.

Chatterjee, M., & Oberzut, C. (2011). Detection and rate discrimination of amplitude modulation in electrical hearing. The Journal of the Acoustical Society of America, 130(3), 1567–1580.

Chatterjee, M., & Peng, S. C. (2008). Processing F0 with cochlear implants: Modulation frequency discrimination and speech intonation recognition. Hearing Research, 235(1–2), 143–156.

Consortium, M. (2020). Quantifying sources of variability in infancy research using the infant-directed speech preference. Advances in Methods and Practices in Psychological Science, 3(1), 24–52. https://doi.org/10.17605/OSF.IO/S98AB

Cooper, R. P., Abraham, J., Berman, S., & Staska, M. (1997). The development of infants’ preference for motherese. Infant Behavior and Development, 20(4), 477–488.

Cooper, R. P., & Aslin, R. N. (1990). Preference for Infant-Directed Speech in the First Month after Birth. Child Development, 61(5), 1584–1595.

Crew, J. D., & Galvin, J. J. (2012). Channel interaction limits melodic pitch perception in simulated cochlear implants. The Journal of the Acoustical Society of America, 132(5), EL429–EL435.

Cristia, A., & Seidl, A. (2014). The hyperarticulation hypothesis of infant-directed speech. Journal of Child Language, 41(4), 935.

Croghan, N. B. H., Duran, S. I., & Smith, Z. M. (2017). Re-examining the relationship between number of cochlear implant channels and maximal speech intelligibility. The Journal of the Acoustical Society of America, 142(6), EL537–EL543.

Cutler, A., Dahan, D., & van Donselaar, W. (1997). Prosody in the Comprehension of Spoken Language: A Literature Review. Language and Speech, 40, 141–201.

Davidson, L. S., Geers, A. E., & Nicholas, J. G. (2014). The effects of audibility and novel word learning ability on vocabulary level in children with cochlear implants. Cochlear Implants International, 15(4), 211–221.

Dilley, L., Lehet, M., Wieland, E. A., Arjmandi, M. K., Houston, D., Kondaurova, M., … Bergeson, T. (2020). Individual differences in mothers’ spontaneous infant-directed speech predict language attainment in children with cochlear implants. Journal of Speech, Language, and Hearing Research, 1–15.

Dorman, M. F., Loizou, P. C., Fitzke, J., & Tu, Z. (1998). The recognition of sentences in noise by normal-hearing listeners using simulations of cochlear-implant signal processors with 6– 20 channels. The Journal of the Acoustical Society of America, 104(6), 3583–3585.

Dorman, M. F., Loizou, P. C., & Rainey, D. (1997a). Simulating the effect of cochlear-implant electrode insertion depth on speech understanding. The Journal of the Acoustical Society of America, 102(5), 2993–2996.

Dorman, M. F., Loizou, P. C., & Rainey, D. (1997b). Speech intelligibility as a function of the number of channels of stimulation for signal processors using sine-wave and noise-band outputs. The Journal of the Acoustical Society of America, 102(4), 2403–2411.

Dunn, C. C., Walker, E. A., Oleson, J., Kenworthy, M., Voorst, T. Van, Tomblin, J. B., … Gantz, B. J. (2014). Longitudinal speech perception and language performance in pediatric cochlear implant users: The effect of age at implantation. Ear and Hearing, 35(2), 148–160.

Eskenazi, M. (1993). Trends in speaking styles research. Third European Conference on Speech Communication and Technology, 501–509.

Falk, T. H., Parsa, V., Santos, J. F., Arehart, K., Hazrati, O., Falk, T. H., … Scollie, S. (2015). Objective Quality Prediction for Users of and Intelligibility Assistive Listening Devices. IEEE Signal Processing Magazine, 32(2), 114–124.

Ferguson, S. H., & Kewley-Port, D. (2007). Talker differences in clear and conversational speech: Acoustic characteristics of vowels. Journal of Speech, Language, and Hearing Research, 50(5), 1241–1255.

Ferguson, S. H., Poore, M. A., Shrivastav, R., Kendrick, A., McGinnis, M., & Perigoe, C. (2010). Acoustic Correlates of Reported Clear Speech Strategies. Journal of the Academy of Rehabilitative Audiology, 43, 45–64.

Ferguson, S. H., & Quené, H. (2014). Acoustic correlates of vowel intelligibility in clear and conversational speech for young normal-hearing and elderly hearing-impaired listeners. The Journal of the Acoustical Society of America, 135(6), 3570–3584.

Fernald, A. (1985). Four-month-old infants prefer to listen to motherese. Infant Behavior and Development, 8(2), 181–195.

Fernald, A. (1989). Intonation and communicative intent in mothers’ speech to infants: Is the melody the message? Child Development, 60(6), 1497–1510.

Fernald, A. (1993). Approval and Disapproval : Infant Responsiveness to Vocal Affect in Familiar and Unfamiliar Languages. Child Development, 64(3), 657–674.

Fernald, A., & Mazzie, C. (1991). Prosody and focus in speech to infants and adults. Developmental Psychology, 27(2), 209–221.

Fernald, A., & Simon, T. (1984). Expanded intonation contours in mothers’ speech to newborns. Developmental Psychology, 20(1), 104–113.

Fernald, & Kuhl. (1987). Acoustic determinants of infant preference for motherse speech. Infant Behaviour and Development, 10, 279–293.

Fetterman, B. L., & Domico, E. H. (2002). Speech recognition in background noise of cochlear implant patients. Otolaryngology - Head and Neck Surgery, 126(3), 257–263.

Friesen, L. M., Shannon, R. V., Baskent, D., & Wang, X. (2001). Speech recognition in noise as a function of the number of spectral channels: Comparison of acoustic hearing and cochlear implants. The Journal of the Acoustical Society of America, 110(2), 1150–1163.

Fu, Q.-J. (2019). AngelSim: Cochlear implant and hearing loss simulator. Retrieved from http://www.tigerspeech.com/angelsim/angelsim_about.html

Fu, Q.-J., Chinchilla, S., & Galvin, J. J. (2004). The role of spectral and temporal cues in voice gender discrimination by normal-hearing listeners and cochlear implant users. Journal of the Association for Research in Otolaryngology, 5(3), 253–260.

Fu, Q.-J., Chinchilla, S., Nogaki, G., & Galvin, J. J. (2005). Voice gender identification by cochlear implant users: The role of spectral and temporal resolution. The Journal of the Acoustical Society of America, 118(3), 1711–1718.

Fu, Q.-J., & Nogaki, G. (2005). Noise susceptibility of cochlear implant users: The role of spectral resolution and smearing. Journal of the Association for Research in Otolaryngology, 6(1), 19–27.

Fu, Q.-J., Shannon, R. V., & Wang, X. (1998). Effects of noise and spectral resolution on vowel and consonant recognition: Acoustic and electric hearing. The Journal of the Acoustical Society of America, 104(6), 3586–3596.

Fuller, C. D., Gaudrain, E., Clarke, J. N., Galvin, J. J., Fu, Q.-J., Free, R. H., & Başkent, D. (2014). Gender Categorization in Cochlear Implant Users. JARO - Journal of the Association for Research in Otolaryngology, 15(6), 1037–1048. https://doi.org/10.1007/s10162-014-0483-7

Garnica, O. (1977). On some prosodic and paralinguistic features of speech to young children. In C. S. and C. Ferguson (Ed.), Talking to Children: Language Input and Acquisition (pp. 271–285). Cambridge, UK: Cambridge University Press.

Gaudrain, E., & Baskent, D. (2018). Discrimination of voice pitch and vocal-tract length in cochlear implant users. Ear and Hearing, 39(2), 226–237. https://doi.org/10.1097/AUD.0000000000000480

Geers, A. E., Strube, M. J., Tobey, E. A., & Moog, J. S. (2011). Epilogue: factors contributing to long-term outcomes of cochlear implantation in early childhood. Ear and Hearing, 32(1 Suppl), 84S.

Geers, Ann E., Nicholas, J., Tobey, E., & Davidsonb, L. (2016). Persistent Language Delay Versus Late Language Emergence in Children With Early Cochlear Implantation. Journal of Speech, Language, and Hearing Research, 59(1), 155–170.

Glenn, S. M., Cunningham, C. C., & Joyce, P. F. (1981). A Study of Auditory Preferences in Nonhandicapped Infants and Infants with down’s Syndrome. Child Development, 1303–1307.

Gogate, L. J., Bahrick, L. E., & Watson, J. D. (2000). A study of multimodal motherese: The role of temporal synchrony between verbal labels and gestures. Child Development, 71(4), 878–894.

Green, J. R., Nip, I. S. B., Wilson, E. M., Mefferd, A. S., & Yunusova, Y. (2010). Lip movement exaggerations during infant-directed speech. Journal of Speech, Language, and Hearing Research, 53(6), 1529–1542.

Green, T., Faulkner, A., & Rosen, S. (2002). Spectral and temporal cues to pitch in noise-excited vocoder simulations of continuous-interleaved-sampling cochlear implants. The Journal of the Acoustical Society of America, 112(5), 2155–2164.

Greenwood, D. D. (1990). A cochlear frequency-position function for several species—29 years later. Journal of the Acoustical Society of America, 87(6), 2592–2605.

Grieco-calub, T. M., Saffran, J. R., & Litovsky, R. Y. (2010). Spoken Word Recognition in Toddlers Who Use Cochlear Implants. Journal of Speech Language and Hearing Research, 52(6), 1390–1400.

Hawkins, S. (2004). Puzzles and patterns in 50 years of research on speech perception. From Sound to Sense: 50+ Years of Discoveries in Speech Communication, 223–246.

Hazan, V., Tuomainen, O., Kim, J., Davis, C., Sheffield, B., & Brungart, D. (2018). Clear speech adaptations in spontaneous speech produced by young and older adults. The Journal of the Acoustical Society of America, 144(3), 1331–1346.

Hazrati, O., & Loizou, P. C. (2012). The combined effects of reverberation and noise on speech intelligibility by cochlear implant listeners. International Journal of Audiology, 51(6), 437–443.

Heijden, V. Der, Ferdinand, R. P. D., Ridder, D. De, & Tax, D. M. (2005). Classification, Parameter Estimation and State Estimation An Engineering Approach Using MATLAB. John Wiley & Sons.

Hoff, E. (2006). How social contexts support and shape language development. Developmental Review, 26(1), 55–88.

Horowitz, F. D. (1983). The Effects of Intonation on Infant Attention: The Role of the Rising Intonation Contour. Journal of Child Language, 10(3), 521–534.

Houston, D. M., Pisoni, D. B., Kirk, K. I., Ying, E. A., & Miyamoto, R. T. (2003). Speech perception skills of deaf infants following cochlear implantation: A first report. International Journal of Pediatric Otorhinolaryngology, 67(5), 479–495.

Houston, D. M., Stewart, J., Moberly, A., Hollich, G., & Miyamoto, R. T. (2012). Word learning in deaf children with cochlear implants: Effects of early auditory experience. Developmental Science, 15(3), 448–461.

Hunt, N. J., Lennig, N., & Mermeletein, P. (1980). Experiments in syllable-based recognition of continuous speech. IEEE International Conference on Acoustics, Speech, and Signal Processing, (5), 880–883.

Imai, S. (1983). Cepstral analysis synthesis on the mel frequency scale. IEEE International Conference on Acoustics, Speech, and Signal Processing, 93–96.

Inoue, T., Nakagawa, R., Kondou, M., Koga, T., & Shinohara, K. (2011). Discrimination between mothers’ infant- and adult-directed speech using hidden Markov models. Neuroscience Research, 70(1), 62–70.

Jahn, K. N., DiNino, M., & Arenberg, J. G. (2019). Reducing Simulated Channel Interaction Reveals Differences in Phoneme Identification Between Children and Adults With Normal Hearing. Ear and Hearing, 40(2), 295–311.

Jain, S., & Vipin Ghosh, P. G. (2018). Acoustic simulation of cochlear implant hearing: Effect of manipulating various acoustic parameters on intelligibility of speech. Cochlear Implants International, 19(1), 46–53.

Janse, E., Nooteboom, S. G., & Quené, H. (2007). Coping with gradient forms of /t/-deletion and lexical ambiguity in spoken word recognition. In Language and Cognitive Processes (Vol. 22).

Jusczyk, P. W., Hohne, E. A., & Bauman, A. (1999). Infants’ sensitivity to allophonic cues for word segmentation. Perception and Psychophysics, 61(8), 1465–1476.

Karzon, R. G. (1985). Discrimination of polysyllabic sequences by one- to four-month-old infants. Journal of Experimental Child Psychology, 39(2), 326–342.

Kirk, K. I., Hay-McCutcheon, M. J., Holt, R. F., Gao, S., Qi, R., & Gerlain, B. L. (2007). Audiovisual spoken word recognition by children with cochlear implants. Audiological Medicine, 5(4), 250–261.

Kirk, K. I., & Pisoni, D. B. (2002). Audiovisual integration of speech by children and adults with cochlear implants. International Conference on Spoken Language Processing, 1689.

Kisilevsky, B. S., Hains, S. M., Lee, K., Xie, X., Huang, H., Ye, H. H., … Wang, Z. (2003). Effects of experience on fetal voice recognition. Psychological Science, 14(3), 220–224.

Kitamura, C., Thanavishuth, C., Burnham, D., & Luksaneeyanawin, S. (2001). Universality and specificity in infant-directed speech: Pitch modifications as a function of infant age and sex in a tonal and non-tonal language. Infant Behavior and Development, 24(4), 372–392.

Kong, Y.-Y., & Carlyon, R. P. (2010). Temporal pitch perception at high rates in cochlear implants. The Journal of the Acoustical Society of America, 127(5), 3114–3123.

Kong, Y. Y., Mullangi, A., Marozeau, J., & Epstein, M. (2011). Temporal and Spectral Cues for Musical Timbre Perception in Electric Hearing. Journal of Speech, Language, and Hearing Research, 54, 981–994. https://doi.org/10.1044/1092-4388(2010/10-0196).Temporal

Kong, Ying Yee, Cruz, R., Jones, J. A., & Zeng, F. G. (2004). Music Perception with Temporal Cues in Acoustic and Electric Hearing. Ear and Hearing, 25(2), 173–185.

Kong, Ying Yee, Stickney, G. S., & Zeng, F.-G. (2005). Speech and melody recognition in binaurally combined acoustic and electric hearing. The Journal of the Acoustical Society of America, 117(3), 1351–1361.

Krause, J. C., & Braida, L. D. (2002). Investigating alternative forms of clear speech: The effects of speaking rate and speaking mode on intelligibility. The Journal of the Acoustical Society of America, 112(5), 2165–2172.

Kuhl, P. K. (2004). Early language acquisition: cracking the speech code. Nature Reviews Neuroscience, 5(11), 831–843.

Kuhl, P. K., Andruski, J. E., Chistovich, I. A., Chistovich, L. A., Kozhevnikova, E. V., Ryskina, L., … Lacerda, F. (1997). Cross-language analysis of phonetic units in language addressed to infants. Science, 277(5326), 684–686.

Kuhl, P. K., & Meltzoff, A. N. (1999). The intermodal representation of speech in newborns. Developmental Science, 2(1), 42–46.

Lachs, L., Pisoni, D. B., & Kirk, K. I. (2001). Use of audiovisual information in speech perception by prelingually deaf children with cochlear implants: A first report. Ear and Hearing, 22(3), 236–251.

Lavan, N., Burton, A. M., Scott, S. K., & McGettigan, C. (2019). Flexible voices: Identity perception from variable vocal signals. Psychonomic Bulletin and Review, 26(1), 90–102.

Leong, V., Kalashnikova, M., Burnham, D., & Goswami, U. (2014). Infant-directed speech enhances temporal rhythmic structure in the envelope. Proceedings of the Annual Conference of the International Speech Communication Association, INTERSPEECH, (September), 2563–2567.

Li, Y., Zhang, G., Kang, H., Liu, S., Han, D., & Fu, Q.-J. (2011). Effects of speaking style on speech intelligibility for Mandarin-speaking cochlear implant users. The Journal of the Acoustical Society of America, 129(6), EL242–EL247.

Litvak, L., Delgutte, B., & Eddington, D. (2001). Auditory nerve fiber responses to electric stimulation: Modulated and unmodulated pulse trains. The Journal of the Acoustical Society of America, 110(1), 368–379.

Liu, H. M., Kuhl, P. K., & Tsao, F. M. (2003). An association between mothers’ speech clarity and infants’ speech discrimination skills. Developmental Science, 6(3), 1–10.

Liu, S., Del Rio, E., Bradlow, A. R., & Zeng, F.-G. (2004). Clear speech perception in acoustic and electric hearing. The Journal of the Acoustical Society of America, 116(4), 2374–2383.

Loizou, P. C., Dorman, M., & Tu, Z. (1999). On the number of channels needed to understand speech. The Journal of the Acoustical Society of America, 106(4), 2097–2103.

Luo, H., & Poeppel, D. (2007). Phase Patterns of Neuronal Responses Reliably Discriminate Speech in Human Auditory Cortex. Neuron, 54(6), 1001–1010.

Luo, X., Fu, Q.-J., & Galvin, J. J. (2007). Vocal Emotion Recognition by Normal-Hearing Listeners and Cochlear Implant Users. Trends in Amplification, 11(4), 301–315.

Ma, W., Golinkoff, R. M., Houston, D. M., & Hirsh-Pasek, K. (2011). Word learning in infant- and adult-directed speech. Language Learning and Development, 7(3), 185–201.

Maesschalck, R. D., & Massart, D. L. (2000). The Mahalanobis distance. Chemometrics and Intelligent Laboratory Systems, 50(1), 1–18.

Masnan, M. J., Mahat, N. I., Shakaff, A. Y. M., Abdullah, A. H., Zakaria, N. Z. I., Yusuf, N., … Aziz, A. H. A. (2015). Understanding Mahalanobis distance criterion for feature selection. AIP Conference Proceedings, 1660, 050075. AIP Publishing LLC.

Mastropieri, D., & Turkewitz, G. (1999). Prenatal experience and neonatal responsiveness to vocal expressions of emotion. Developmental Psychobiology, 35(3), 204–214.

Mattys, S. L. (2000). The perception of primary and secondary stress in English. Perception and Psychophysics, 62(2), 253–265.

Matuschek, H., Kliegl, R., Vasishth, S., Baayen, H., & Bates, D. (2017). Balancing Type I error and power in linear mixed models. Journal of Memory and Language, 94, 305–315.

Maye, J., Werker, J. F., & Gerken, L. A. (2002). Infant sensitivity to distributional information can affect phonetic discrimination. Cognition, 82(3), 101–111.

McMurray, B., & Aslin, R. N. (2005). Infants are sensitive to within-category variation in speech perception. Cognition, 95(2), B15–B26.

McMurray, B., Farris-Trimble, A., & Rigler, H. (2017). Waiting for lexical access: Cochlear implants or severely degraded input lead listeners to process speech less incrementally. Cognition, 169(August), 147–164.

McMurray, B., Kovack-Lesh, K. A., Goodwin, D., & McEchron, W. (2013). Infant directed speech and the development of speech perception: Enhancing development or an unintended consequence? Cognition, 85(0 1), 1–27.

Mehler, J. (1981). The role of syllables in speech processing: infant and adult data. Philosophical Transactions of the Royal Society of London. B, Biological Sciences, 295(1077), 333–352.

Mehta, A. H., Lu, H., & Oxenham, A. J. (2020). The Perception of Multiple Simultaneous Pitches as a Function of Number of Spectral Channels and Spectral Spread in a Noise-Excited Envelope Vocoder. JARO - Journal of the Association for Research in Otolaryngology, 21(1), 61–72.

Mehta, A. H., & Oxenham, A. J. (2017). Vocoder Simulations Explain Complex Pitch Perception Limitations Experienced by Cochlear Implant Users. JARO - Journal of the Association for Research in Otolaryngology, 18(6), 789–802.

Miller, J. L. (1994). On the internal structure of phonetic categories: a progress report. Cognition, 50(1–3), 271–285.

Miyamoto, R. T., Houston, D. M., Kirk, K. I., Perdew, A. E., & Svirsky, M. A. (2003). Language development in deaf infants following cochlear implantation. Acta Oto-Laryngologica, 123(2), 241–244.

Moore, B. C. J. (2003). Coding of sounds in the auditory system and its relevance to signal processing and coding in cochlear implants. Otology and Neurotology, 24(2), 243–254.

Moore, D. S., Spence, M. J., & Katz, G. S. (1997). Six-month-olds’ categorization of natural infant-directed utterances. Developmental Psychology, 33(6), 980–989.

Nagels, L., Bastiaanse, R., Başkent, D., & Wagner, A. (2020). Individual differences in lexical access among cochlear implant users. Journal of Speech, Language, and Hearing Research, 63(1), 286–304.

Narayan, C. R., & McDermott, L. C. (2016). Speech rate and pitch characteristics of infant-directed speech: Longitudinal and cross-linguistic observations. The Journal of the Acoustical Society of America, 139(3), 1272–1281.

Niparko, J. K., Tobey, E. A., Thal, D. J., Eisenberg, L. S., Wang, N.-Y., Quittner, A. L., & Fink, N. E. (2010). Spoken language development in children following cochlear implantation. Jama, 303(15), 1498–1506.

Nomikou, I., & Rohlfing, K. J. (2011). Language does something: Body action and language in maternal input to three-month-olds. IEEE Transactions on Autonomous Mental Development, 3(2), 113–128.

Papoušek, M., Bornstein, M. H., Nuzzo, C., Papoušek, H., & Symmes, D. (1990). Infant responses to prototypical melodic contours in parental speech. Infant Behavior and Development, 13(4), 539–545.

Peng, S. C., Lu, H. P., Lu, N., Lin, Y. S., Deroche, M. L. D., & Chatterjee, M. (2017). Processing of acoustic cues in lexical-tone identification by pediatric cochlear-implant recipients. Journal of Speech, Language, and Hearing Research, 60(5), 1223–1235.

Peng, S. C., Tomblin, J. B., & Turner, C. (2008). Production and Perception of Speech Intonation in Pediatric Cochlear Implant Recipients and Individuals with Normal Hearing. Ear and Hearing, 29(3), 336–351.

Peng, Z., Hess, C., Saffran, J. R., Edwards, J. R., & Litovsky, R. Y. (2019). Assessing Fine-Grained Speech Discrimination in Young Children With Bilateral Cochlear Implants. Otology & Neurotology, 40(3), e191–e197.

Peter, J. W., & Robert S S. (2008). The effect of fundamental frequency on the intelligibility of speech with flattened intonation contours. American Journal of Speech-Language Pathology, 17(4), 348–355.

Piazza, E. A., Iordan, M. C., & Lew-Williams, C. (2017). Mothers Consistently Alter Their Unique Vocal Fingerprints When Communicating with Infants. Current Biology, 27(20), 3162- 3167.e3.

Pisoni, D. B., Conway, C. M., Kronenberger, W., Horn, D. L., Karpicke, J., & Henning, S. (2007). Efficacy and effectiveness of cochlear implants in deaf children. Research on Spoken Language Processing, 28(28), 3–46.

Pisoni, D. B., Kronenberger, W. G., Harris, M. S., & Moberly, A. C. (2018). Three challenges for future research on cochlear implants. World Journal of Otorhinolaryngology - Head and Neck Surgery, 3(4), 240–254.

Qin, M. K., & Oxenham, A. J. (2003). Effects of simulated cochlear-implant processing on speech reception in fluctuating maskers. The Journal of the Acoustical Society of America, 114(1), 446–454.

Qin, M. K., & Oxenham, A. J. (2005). Effects of Envelope-Vocoder Processing on F0 Discrimination. Ear & Hearing, 26(5), 451–460.

Quené, H., & van den Bergh, H. (2008). Examples of mixed-effects modeling with crossed random effects and with binomial data. Journal of Memory and Language, 59(4), 413–425.

Rohlfing, K. J., Fritsch, J., Wrede, B., & Jungmann, T. (2006). How can multimodal cues from child-directed interaction reduce learning complexity in robots? Advanced Robotics, 20(10), 1183–1199.

Rosen, S., Faulkner, A., & Wilkinson, L. (1999). Adaptation by normal listeners to upward spectral shifts of speech: Implications for cochlear implants. The Journal of the Acoustical Society of America, 106(6), 3629–3636.

Rottmann, N., & Zobrist, P. (2004). Tuned to the signal: The privileged status of speech for young infants. Developmental Science, 7(3), 270–276.

Rubinstein, J. T., Wilson, B. S., Finley, C. C., & Abbas, P. J. (1999). Pseudospontaneous activity: Stochastic independence of auditory nerve fibers with electrical stimulation. Hearing Research, 127(1–2), 108–118.

Rubinstein, Jay T. (2004). How cochlear implants encode speech. Current Opinion in Otolaryngology and Head and Neck Surgery, 12(5), 444–448.

Santos, J. F., Cosentino, S., Hazrati, O., Loizou, P. C., & Falk, T. H. (2013). Objective speech intelligibility measurement for cochlear implant users in complex listening environments. Speech Communication, 55(7–8), 815–824.

Sato, N., & Obuchi, Y. (2007). Emotion Recognition using Mel-Frequency Cepstral Coefficients. Information and Media Technologies, 2(3), 835–848.

Schachner, A., & Hannon, E. E. (2011). Infant-Directed Speech Drives Social Preferences in 5- Month-Old Infants. Developmental Psychology, 47(1), 19–25.

Shaneh, M., & Taheri, A. (2009). Voice command recognition system based on MFCC and VQ algorithms. World Academy of Science, Engineering and Technology, 57, 534–538.

Shannon, R. V., Zeng, F.-G., Kamath, V., Wygonski, J., & Ekelid, M. (1995). Speech recognition with primarily temporal cues. Science, 270(5234), 303–304.

Singh, L. (2008). Influences of high and low variability on infant word recognition. Cognition, 106(2), 833–870.

Singh, L., Morgan, J. L., & Best, C. T. (2002). Infants’ listening preferences: Baby talk or happy talk? Infancy, 3(3), 365–394.

Singh, L., & Nestor, S. (2009). Influences of Infant-Directed Speech on Early Word Recognition. Infancy, 14(6), 654–666.

Slaney, M., & Mcroberts, G. (1998). BABY EARS: A recognition system for affective vocalizations. IEEE International Conference on Acoustics, Speech and Signal Processing, 985–988.

Snow, C. E. (1977). Mothers’ speech research: From input to interaction. Talking to Children: Language Input and Acquisition, 3149.

Sommers, M. S., Nygaard, L. C., & Pisoni, D. B. (1992). Stimulus variability and the perception of spoken words: Effects of variations in speaking rate and overall amplitude. International Conference on Spoken Language Processing, (October), 217–220.

Song, J. Y., Demuth, K., & Morgan, J. (2010). Effects of the acoustic properties of infant-directed speech on infant word recognition. The Journal of the Acoustical Society of America, 128(1), 389–400.

Souza, P., Gehani, N., Wright, R., & McCloy, D. (2013). The advantage of knowing the talker. Journal of the American Academy of Audiology, 24(8), 689–700.

Spencer, P. E. (2004). Individual Differences in Language Performance after Cochlear Implantation at One to Three Years of Age: Child, Family, and Linguistic Factors. Journal of Deaf Studies and Deaf Education, 9(4), 395–412.

Spitzer, S. M., Liss, J. M., & Mattys, S. L. (2007). Acoustic cues to lexical segmentation: A study of resynthesized speech. The Journal of the Acoustical Society of America, 122(6), 3678–3687.

Spitzer, S. M., Liss, J., Spahr, T., Dorman, M., & Lansford, K. (2009). The use of fundamental frequency for lexical segmentation in listeners with cochlear implants. The Journal of the Acoustical Society of America, 125(6), EL236–EL241.

Sulpizio, S., Kuroda, K., Dalsasso, M., Asakawa, T., Bornstein, M. H., Doi, H., … Shinohara, K. (2018). Discriminating between Mothers’ Infant- and Adult-Directed Speech: Cross-Linguistic Generalizability from Japanese to Italian and German. Neurosci Res., 133, 21–27.

Svirsky, M. A. (2017). Cochlear implants and electronic hearing. Physics Today, 70(8), 53–58.

Szagun, G., & Schramm, S. A. (2016). Sources of variability in language development of children with cochlear implants: Age at implantation, parental language, and early features of children’s language construction. Journal of Child Language, 43(3), 505–536.

Tamati, T. N., Janse, E., & Başkent, D. (2019). Perceptual Discrimination of Speaking Style under Cochlear Implant Simulation. Ear and Hearing, 40(1), 63–76.

Telkemeyer, S., Rossi, S., Koch, S. P., Nierhaus, T., Steinbrink, J., Poeppel, D., … Wartenburger, I. (2009). Sensitivity of Newborn Auditory Cortex to the Temporal Structure of Sounds. Journal of Neuroscience, 29(47), 14726–14733.

Thiessen, E. D., Hill, E. A., & Saffran, J. R. (2005). Infant directed speech facilitates word segmentation. Infancy, 7(1), 53–71.

Throckmorton, C. S., & Collins, L. M. (2002). The effect of channel interactions on speech recognition in cochlear implant subjects: Predictions from an acoustic model. The Journal of the Acoustical Society of America, 112(1), 285–296.

Trainor, L. J., Austin, C. M., & Desjardins, N. (2000). Is infant-directed speech prosody a result of the vocal expression of emotion? Psychological Science, 11(3), 188–195.

Walker-Andrews A. S. & Grolnick, W. (1983). Discrimination of vocal expression by young infants. Infant Behavior and Development, 6, 491–498.

Walker-Andrews, A. S., & Lennon, E. (1991). Infants’ discrimination of vocal expressions: Contributions of auditory and visual information. Infant Behavior and Development, 14(2), 131–142.

Wang, Y., Bergeson, T. R., & Houston, D. M. (2017). Infant-Directed Speech Enhances Attention to Speech in Deaf Infants With Cochlear Implants. Journal of Speech Language and Hearing Research, 60(11), 3321.

Wang, Y., Bergeson, T. R., & Houston, D. M. (2018). Preference for Infant-Directed Speech in Infants With Hearing Aids: Effects of Early Auditory Experience. Journal of Speech, Language & Hearing Research, 61(9), 2431–2439.

Wang, Y., Shafto, C. L., & Houston, D. M. (2018). Attention to speech and spoken language development in deaf children with cochlear implants: a 10-year longitudinal study. Developmental Science, (July 2017), e12677.

Webb, A. R., Heller, H. T., Benson, C. B., & Lahav, A. (2015). Mother’s voice and heartbeat sounds elicit auditory plasticity in the human brain before full gestation. Proceedings of the National Academy of Sciences of the United States of America, 112(10), 3152–3157.

Weisleder, A., & Fernald, A. (2013). Talking to Children Matters: Early Language Experience Strengthens Processing and Builds Vocabulary. Psychological Science, 24(11), 2143–2152.

Werker, J. F., & McLeod, P. J. (1989). Infant preference for both male and female infant-directed talk: a developmental study of attentional and affective responsiveness. Canadian Journal of Psychology, 43(2), 230–246.

Werker, J. F., Pegg, J., & McLeod, P. J. (1994). A cross-language investigation of infant preference for infant-directed communication. Infant Behavior and Development, 17(3), 323–333.

Winn, M. B., & Litovsky, R. Y. (2015). Using speech sounds to test functional spectral resolution in listeners with cochlear implants. The Journal of the Acoustical Society of America, 137(3), 1430–1442.

Xiang, S., Nie, F., & Zhang, C. (2008). Learning a Mahalanobis distance metric for data clustering and classification. Pattern Recognition, 41(12), 3600–3612.

Zanto, T. P., Hennigan, K., Östberg, M., Clapp, W. C., & Gazzaley, A. (2013). Effect of speaking rate on recognition of synthetic and natural speech by normal-hearing and cochlear implant listeners. Ear and Hearing, 34(3), 313.

Zeng, F. G., Tang, Q., & Lu, T. (2014). Abnormal pitch perception produced by cochlear implant stimulation. PLoS ONE, 9(2).

Zheng, Y., Koehnke, J., & Besing, J. (2011). Effects of Noise and Reverberation on Virtual Sound Localization for Listeners With Bilateral Cochlear Implants. American Journal of Audiology, 32(5), 569–572.

